# The geospatial phylodynamics of SARS-CoV-2 in Nepal and clinical phenotype of long COVID during the Delta-Omicron waves

**DOI:** 10.1101/2025.07.24.25332179

**Authors:** Wytamma Wirth, Tilda N. Thomson, Prajwol Manandhar, Swastika Shrestha, Rajindra Napit, Sameer Mani Dixit, Gyanendra Shrestha, Courtney R. Lane, Pradip Gyanwali, Suman Pant, Sagar Kumar Rajbhandari, Parmananda Bhandari, Bimal Sharma Chalise, Chuman Lal Das, Pranav Kumar Yadav, Prakash Thapa, Prasanna Mishra, Buddha Basnyat, Abhilasha Karkey, Nhukesh Maharjan, Ganna Kovalenko, Ian Goodfellow, Stephen Baker, Lisa J. Ioannidis, Torsten Seemann, Sarah Dunstan, Dipendra Pandey, Reshu Parajuli, Ram Prasad Koirala, Raghu Dhital, Maxine Caws

**Affiliations:** Department of Microbiology and Immunology, University of Melbourne, at The Peter Doherty Institute for Infection and Immunity, Victoria, Australia; Centre for Pathogen Genomics, University of Melbourne, Victoria, Australia; Center for Molecular Dynamics Nepal (CMDN), Kathmandu, Nepal; Birat Nepal Medical Trust (BNMT), Kathmandu, Nepal; Nepal Health Research Council, Kathmandu, Nepal; Sukraraj Tropical and Infectious Disease Hospital, Teku, Kathmandu, Nepal; Koshi Hospital, Biratnagar, Nepal; Bheri Hospital, Nepalgunj, Nepal; Oxford University Clinical Research Unit Nepal; Division of Virology, Department of Pathology, University of Cambridge, Cambridge, UK; ASTAR Infectious Diseases Labs (ASTAR IDL), Agency for Science, Technology and Research (A*STAR), Singapore; Department of Infectious Diseases, University of Melbourne, at The Peter Doherty Institute for Infection and Immunity, Victoria, Australia; Department of Clinical Sciences, Liverpool School of Tropical Medicine, Liverpool, UK

## Abstract

The COVID-19 pandemic exposed gaps in global emerging infectious disease preparedness, particularly for countries with high levels of economic migration, such as Nepal. The Epidemic Intelligence project analysed SARS-CoV-2 genomic and epidemiological data from 2,046 COVID-19 patients across three Nepali regions to study viral dynamics, phylogeography, and long COVID.

Participants were interviewed at diagnosis, and at 3-, 6-, and 12-months post-infection. Long COVID was explored using three standardised definitions from the literature, and logistic regression was used to identify risk factors, stratified by SARS-CoV-2 variant. Over 40% of participants reported long COVID at 3 months defined by symptoms ≥12 weeks and the presence of at least one symptom from the Wellcome Trust Longitudinal Population Study questionnaire. 10.6% (n=211/1992) of participants reported at least two of the symptom clusters from the WHO Delphi consensus definition at 3 months, and 0.7% (n=14/1992) reported all three symptom clusters. The prevalence of long COVID was higher among people infected with the Delta variant than the Omicron variant across all definitions and time points. Among Delta cases, female sex, chronic comorbidity, and full vaccination were associated with long COVID; whereas for people infected with Omicron, only age between 40-54 was a risk factor.

Phylogeographic analyses revealed the epidemiological importance of Nepal’s porous border with India and the changing role of migration in viral spread as the pandemic evolved. Our findings also highlight the burden of post-COVID syndrome in low-resource settings and the need for further research to mitigate the financial and clinical impact. Integration of pathogen genomic analysis with clinical and epidemiological data can guide public health responses during the emergence of novel infectious diseases.

## Introduction

The emergence of COVID-19 in early 2020 and the resulting pandemic highlighted many gaps in global preparedness and response to emerging infectious diseases. On 11 March 2020 the World Health Organisation (WHO) officially declared a pandemic, precipitating a chaotic and often uncoordinated response from countries scrambling to protect their citizens from the largely unknown threat (1, 2). Travel restrictions and lockdowns, often controversial, formed the backbone of the response in many countries (3). Pathogen genomic sequencing provided a valuable tool enabling public health authorities to monitor and track the emergence of new variants of the causative virus, SARS CoV-2, and correlate these with changes in clinical phenotype (4). In early 2021, WHO began to designate emerging variants as either ‘Variants of Interest’ or ‘Variants of Concern’ to classify novel lineages of SARS CoV-2 which were potentially linked to more severe clinical phenotypes, reduced vaccine efficacy or increased transmission potential (5, 6). The definition of such variants evolved during the pandemic.

Despite the unprecedented travel restrictions and lockdowns, emerging SARS CoV-2 variants appeared to rapidly disseminate globally throughout the pandemic. It is important to understand how the novel variants seeded and disseminated in different locations to inform future pandemic preparedness and response strategies.

Nepal is a country of 30 million people which lies in the Himalaya between the two giants of India and China. In recent decades, economic labour migration has surged. Nepal has a remittance ratio exceeding 25% of gross domestic product (GDP), the fifth highest remittance-to-GDP ratio globally, reflecting its heavy reliance on migrant workers (7). Domestically, migrants travel in search of improved livelihoods from rural to urban areas and from mountain/hill districts to the fertile plains region. Internationally, migrants travel to work in diverse sectors including skilled and unskilled labour for hospitality, security, construction, information technology, domestic work, healthcare and domestic caregiving services. Major destination countries are India (through an open border), the Middle East (Qatar, Saudia Arabia, United Arab Emirates), and Malaysia (8). At the time of the pandemic declaration by WHO in March 2020, many Nepali economic migrants were suddenly without livelihoods and forced to return to Nepal, or to return within Nepal to their rural districts of origin: an unprecedented ‘reverse migration’ wave. There was a greater than 10-fold drop in the number of labour permits issued to migrant workers during 2019/2020, compared to the previous 2018/2019 period (9, 10). As the situation evolved, outward economic migration again began to take place both nationally and internationally. Figure 1 gives an overview of the COVID-19 pandemic trajectory in Nepal, showing the timeline of notified cases and deaths from COVID-19 and the major government-imposed lockdowns.

**Figure 1:**
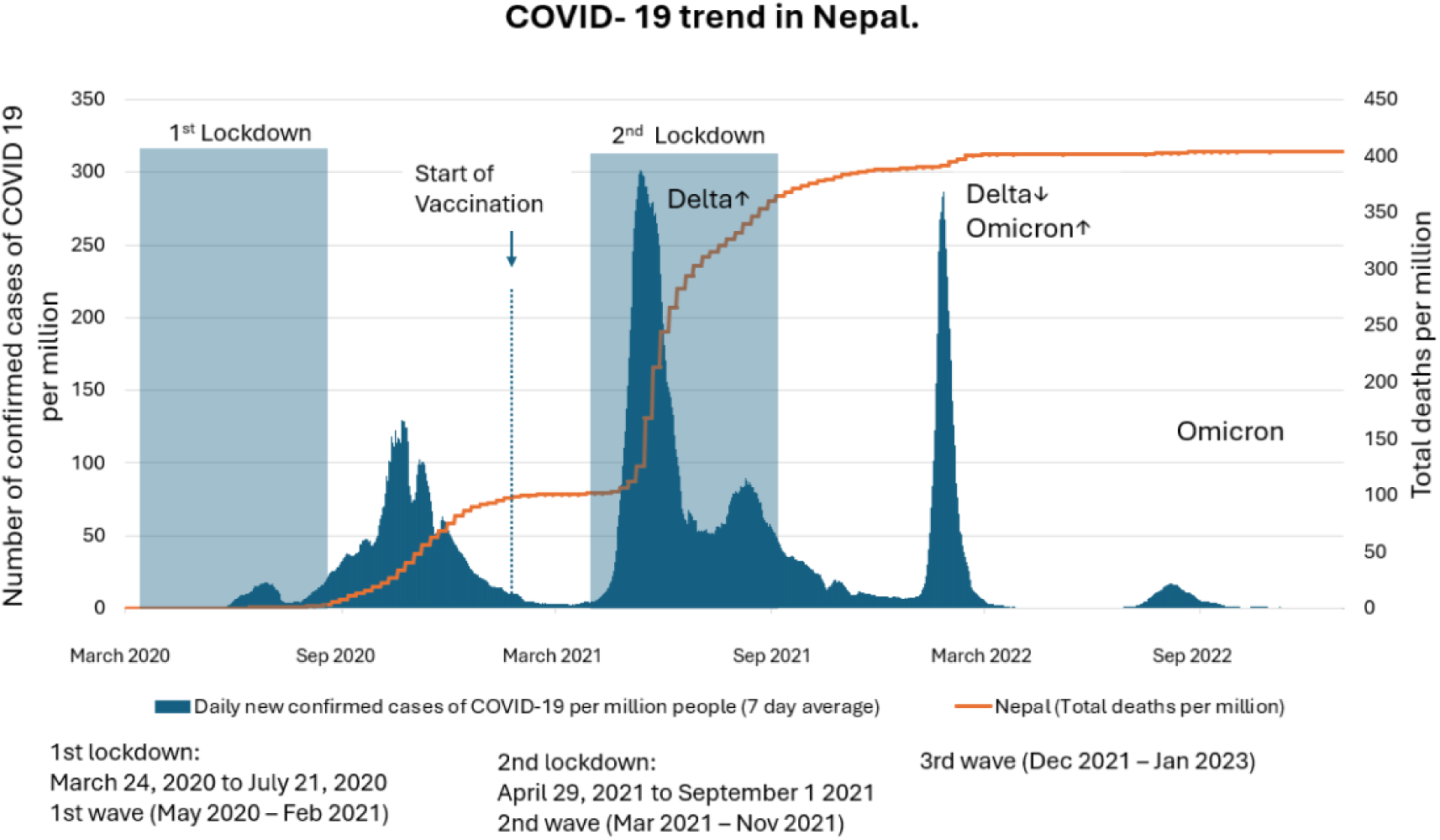
Overview of the COVID-19 epidemic in Nepal showing timeline of case notifications, deaths, major SARS CoV-2 variant waves and phases of major lockdowns. Epidemic Intelligence participants were recruited between June 2021 and February 2023.

In response to the COVID-19 pandemic, the Epidemic Intelligence project (www.epiintelnepal.org) aimed to build capacity for pathogen genome sequencing in Nepal, monitor the introduction and transmission, or *de novo* emergence, of novel SARS CoV-2 variants within Nepal, and investigate the association between variants and epidemiology of COVID-19 and long COVID. We also sought to understand the influence of vaccination and SARS-CoV-2 variants on long COVID risk factors and clinical phenotype.

By analysing SARS-CoV-2 sequence data and epidemiological data collected longitudinally at 0-, 3-, 6- and 12-months following infection, we aimed to describe the phylodynamics and phylogeography of SARS-CoV-2 in Nepal, the prevalence of self-reported long COVID, and to explore factors associated with long COVID, including SARS-CoV-2 genetic variation.

## Results

### Study population

We recruited 2,046 participants with COVID-19 from Koshi (n=780/2046; 38.0%), Bheri (n=678/2046; 33.0%), and Teku (n=588/2046; 29.0%) hospitals. Table 1 summarises the characteristics of the study population, stratified by recruitment location. Participants lived across 11 districts in Nepal, although the majority (n=1,689/2,046; 82.6%) were resident in the districts where the recruiting hospitals are located: Koshi hospital in Morang (n=763; 37.3%), Bheri hospital in Banke (n=460; 22.4%) and Teku hospital in Kathmandu (n=466; 22.8%).

**Table 1:**
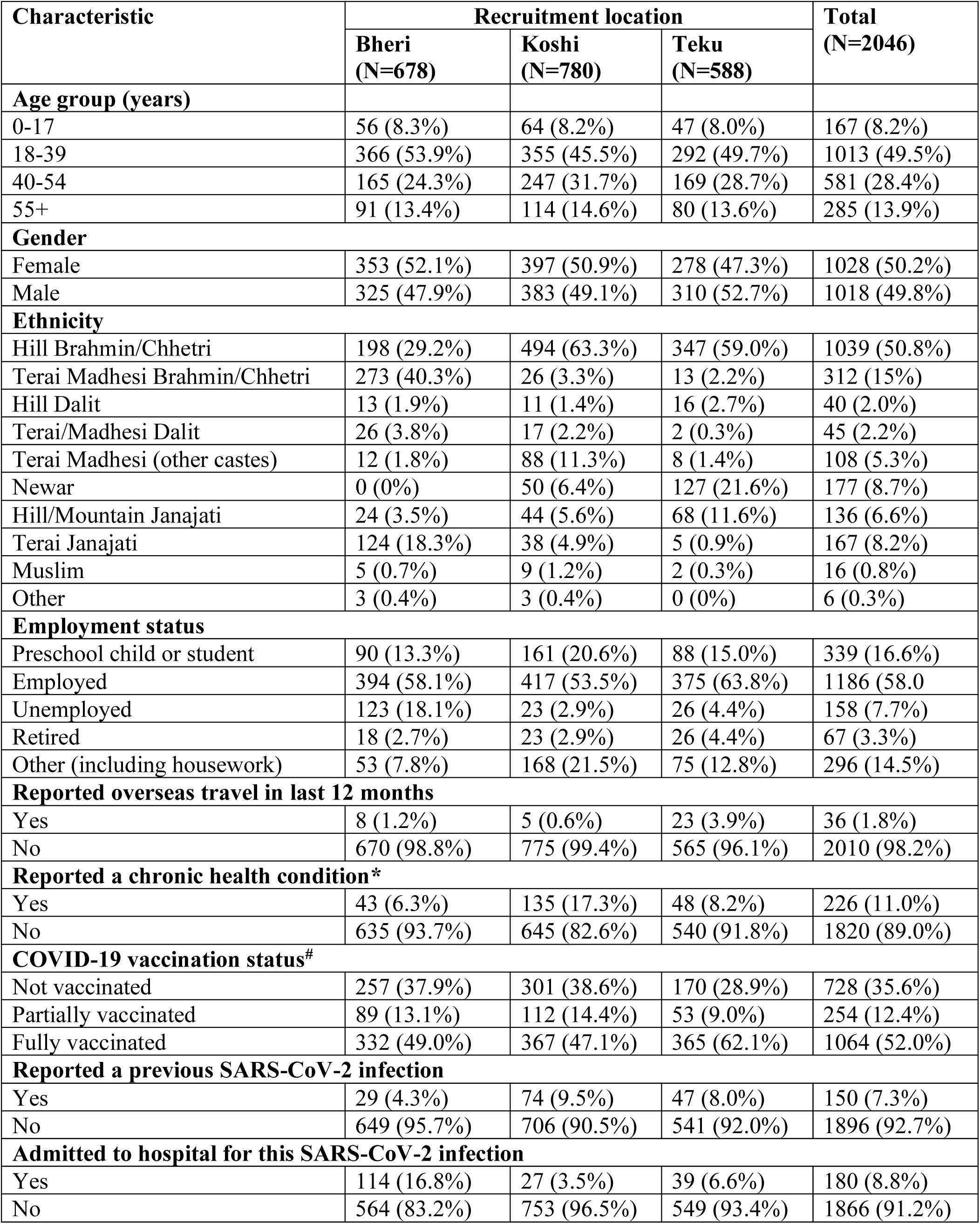
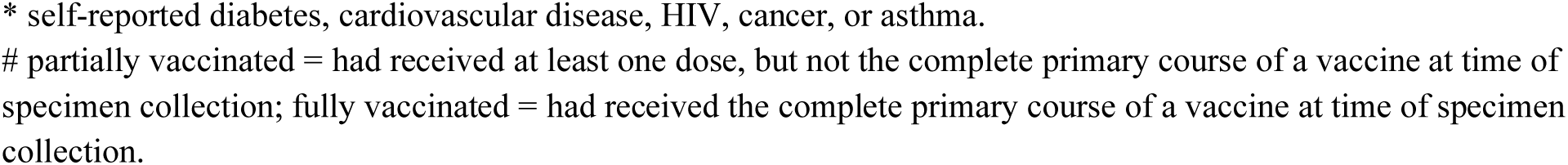
Characteristics of 2,046 participants with COVID-19 who completed the baseline questionnaire.

| Characteristic | Recruitment location |  |  | Total<br>(N=2046) |
| --- | --- | --- | --- | --- |
|  | Bheri<br>(N=678) | Koshi<br>(N=780) | Teku<br>(N=588) |  |
| Age group (years) |  |  |  |  |
| 0-17 | 56 (8.3%) | 64 (8.2%) | 47 (8.0%) | 167 (8.2%) |
| 18-39 | 366 (53.9%) | 355 (45.5%) | 292 (49.7%) | 1013 (49.5%) |
| 40-54 | 165 (24.3%) | 247 (31.7%) | 169 (28.7%) | 581 (28.4%) |
| 55+ | 91 (13.4%) | 114 (14.6%) | 80 (13.6%) | 285 (13.9%) |
| Gender |  |  |  |  |
| Female | 353 (52.1%) | 397 (50.9%) | 278 (47.3%) | 1028 (50.2%) |
| Male | 325 (47.9%) | 383 (49.1%) | 310 (52.7%) | 1018 (49.8%) |
| Ethnicity |  |  |  |  |
| Hill Brahmin/Chhetri | 198 (29.2%) | 494 (63.3%) | 347 (59.0%) | 1039 (50.8%) |
| Terai Madhesi Brahmin/Chhetri | 273 (40.3%) | 26 (3.3%) | 13 (2.2%) | 312 (15%) |
| Hill Dalit | 13 (1.9%) | 11 (1.4%) | 16 (2.7%) | 40 (2.0%) |
| Terai/Madhesi Dalit | 26 (3.8%) | 17 (2.2%) | 2 (0.3%) | 45 (2.2%) |
| Terai Madhesi (other castes) | 12 (1.8%) | 88 (11.3%) | 8 (1.4%) | 108 (5.3%) |
| Newar | 0 (0%) | 50 (6.4%) | 127 (21.6%) | 177 (8.7%) |
| Hill/Mountain Janajati | 24 (3.5%) | 44 (5.6%) | 68 (11.6%) | 136 (6.6%) |
| Terai Janajati | 124 (18.3%) | 38 (4.9%) | 5 (0.9%) | 167 (8.2%) |
| Muslim | 5 (0.7%) | 9 (1.2%) | 2 (0.3%) | 16 (0.8%) |
| Other | 3 (0.4%) | 3 (0.4%) | 0 (0%) | 6 (0.3%) |
| Employment status |  |  |  |  |
| Preschool child or student | 90 (13.3%) | 161 (20.6%) | 88 (15.0%) | 339 (16.6%) |
| Employed | 394 (58.1%) | 417 (53.5%) | 375 (63.8%) | 1186 (58.0) |
| Unemployed | 123 (18.1%) | 23 (2.9%) | 26 (4.4%) | 158 (7.7%) |
| Retired | 18 (2.7%) | 23 (2.9%) | 26 (4.4%) | 67 (3.3%) |
| Other (including housework) | 53 (7.8%) | 168 (21.5%) | 75 (12.8%) | 296 (14.5%) |
| Reported overseas travel in last 12 months |  |  |  |  |
| Yes | 8 (1.2%) | 5 (0.6%) | 23 (3.9%) | 36 (1.8%) |
| No | 670 (98.8%) | 775 (99.4%) | 565 (96.1%) | 2010 (98.2%) |
| Reported a chronic health condition* |  |  |  |  |
| Yes | 43 (6.3%) | 135 (17.3%) | 48 (8.2%) | 226 (11.0%) |
| No | 635 (93.7%) | 645 (82.6%) | 540 (91.8%) | 1820 (89.0%) |
| COVID-19 vaccination status# |  |  |  |  |
| Not vaccinated | 257 (37.9%) | 301 (38.6%) | 170 (28.9%) | 728 (35.6%) |
| Partially vaccinated | 89 (13.1%) | 112 (14.4%) | 53 (9.0%) | 254 (12.4%) |
| Fully vaccinated | 332 (49.0%) | 367 (47.1%) | 365 (62.1%) | 1064 (52.0%) |
| Reported a previous SARS-CoV-2 infection |  |  |  |  |
| Yes | 29 (4.3%) | 74 (9.5%) | 47 (8.0%) | 150 (7.3%) |
| No | 649 (95.7%) | 706 (90.5%) | 541 (92.0%) | 1896 (92.7%) |
| Admitted to hospital for this SARS-CoV-2 infection |  |  |  |  |
| Yes | 114 (16.8%) | 27 (3.5%) | 39 (6.6%) | 180 (8.8%) |
| No | 564 (83.2%) | 753 (96.5%) | 549 (93.4%) | 1866 (91.2%) |

\* self-reported diabetes, cardiovascular disease, HIV, cancer, or asthma.

Half of the participants (n=1,064/2046; 52.0%) were fully vaccinated at the time of infection with SARS-CoV-2 and recruitment into the study (Table 1).

Very few participants (n=36/2046; 1.8%) reported overseas travel in the 12-months preceding their SARS-CoV-2 infection. The most common country visited among those who reported overseas travel was India (n=26/36; 72.2%).

The most common COVID-19 symptoms reported in the baseline questionnaire were fever (n=1572/2046; 76.8%), weakness/tiredness (n=1412/2046; 69.0%) and muscle aches (n=1204/2046; 58.8%) (see Supplementary Materials Figure S1).

### SARS-CoV-2 lineages

From the 2,046 participants, there were 1,764 SARS-CoV-2 sequenced samples in the dataset (see Supplementary Materials Figure S2). The average coverage of the genomes were 96% (std 0.09). Overall, the most common Pango lineage in the dataset was B.1.617.2 (Delta). From 2021 to 2022 the Delta variant dominated, in early 2022 Omicron rapidly displaced Delta, with little overlap between the two major variants. There were more participants infected with the Delta variant, however fewer sublineages were identified among the samples than were identified for Omicron, which is also seen in the global SARS-CoV-2 data. The Epidemic Intelligence study dataset contained 12 distinct Delta and 33 Omicron sublineages. All samples matched previously identified lineages i.e. the first Epidemic Intelligence collection date was after the first collection date for all matching lineages in GISAID. Following collapse of the lineages to either B.1.617.2 (Delta) or B.1.1.529 (Omicron) and removal of outliers (e.g. recombinants), 1193 Delta and 564 Omicron samples remained. Details of the sublineages identified are given in Supplementary Materials Table S1.

### Factors associated with disease severity

Of the 2,046 participants recruited, 180 (8.8%) were admitted to hospital during their illness. Among participants admitted to hospital, 35 (19.4%) were admitted to intensive care and four (0.6%) reported receiving invasive ventilation. The majority of the hospitalised participants were infected with the Delta variant (n=128/180; 71.1%), while few were infected with the Omicron variant (n=21/180; 11.7%). SARS-CoV-2 lineage was undetermined for 31 (17.2%) of the hospitalised participants due to inadequate viral load for sequencing.

Age greater than 55 years was associated with hospital admission for both individuals infected with the Delta (adjusted OR=3.52 [95% CI 2.02, 6.05]; P=<0.001) and Omicron (adjusted OR=5.08 [95% CI 1.32, 18.8]; P=0.015) variants (Table 2). A fully completed primary course of vaccination was protective against hospital admission for those infected with the Delta variant (adjusted OR=0.40 [95% CI 0.25, 0.64]; P<0.001) but showed no association with hospital admission for the Omicron variant (adjusted OR=1.36 [95% CI 0.24, 25.0); P=0.8). The majority of recruited individuals were fully vaccinated by the time of widespread transmission of the Omicron variant. Partial vaccination status (at least one dose of vaccine without completing the full course) did not show any association with hospitalisation for either group. Gender was not a risk factor for hospitalisation for those infected with the Delta variant (adjusted OR=1.38 [95% CI 0.95, 2.03]; P=0.10) but those of female gender were at higher risk of hospitalisation for the Omicron variant (adjusted OR=3.17 [95% CI 1.11, 10.6]; P=0.041).

**Table 2:**
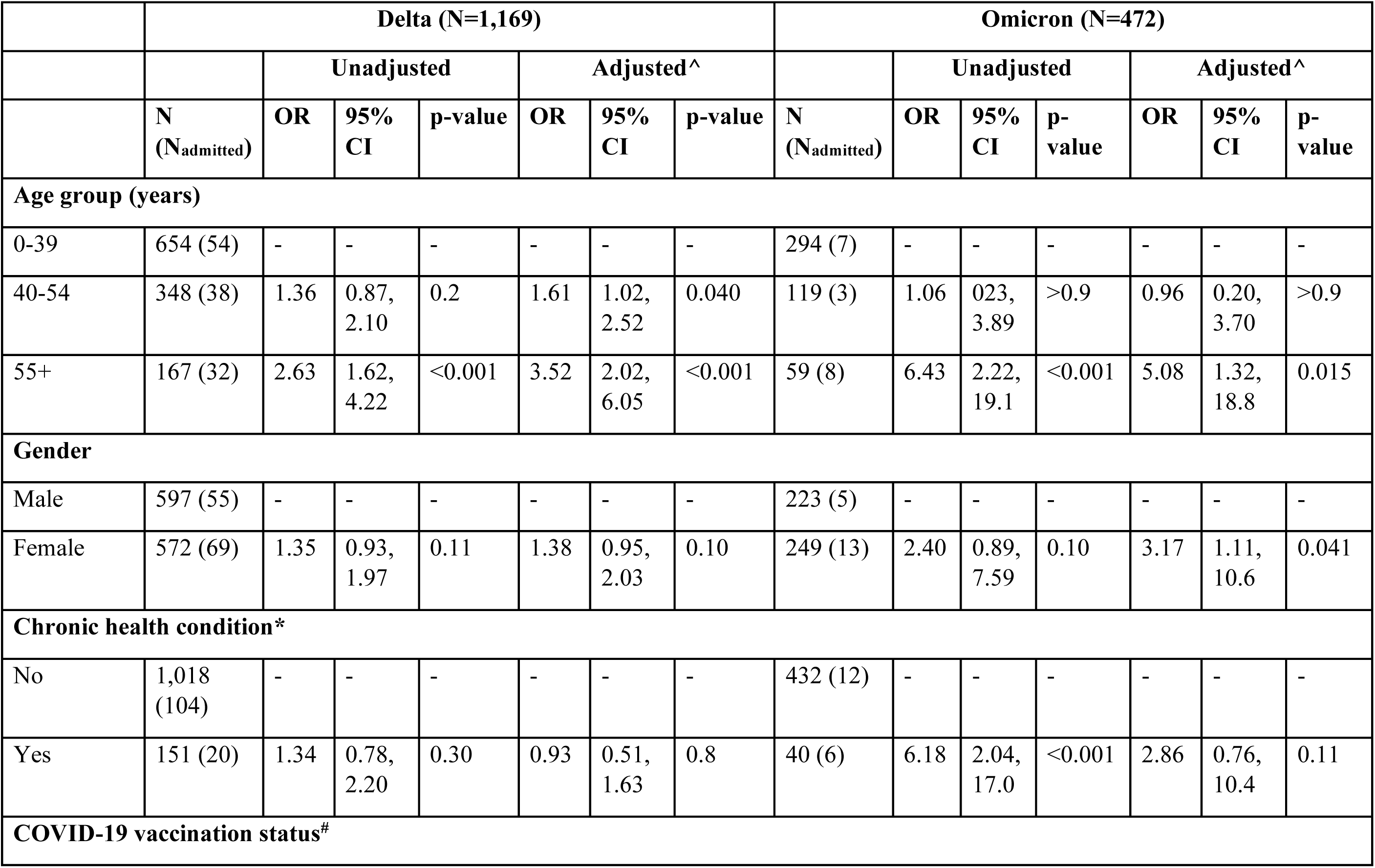
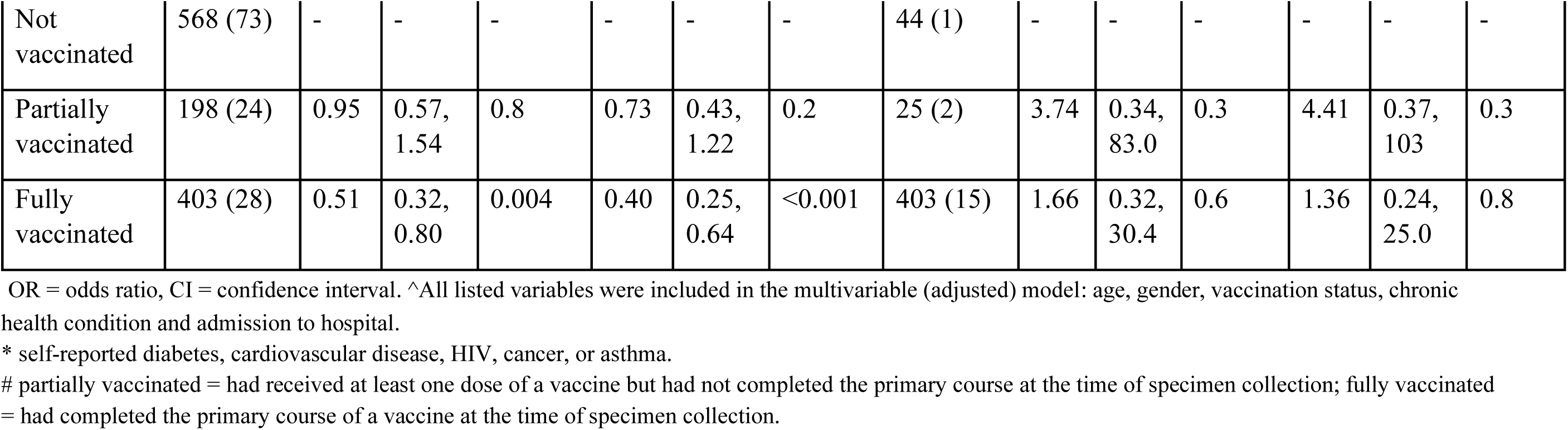
Factors associated with hospital admission, for those infected with Delta and Omicron variants who did not report a previous SARS-CoV-2 infection.

|  | Delta (N=1,169) |  |  |  |  |  |  | Omicron (N=472) |  |  |  |  |  |  |
| --- | --- | --- | --- | --- | --- | --- | --- | --- | --- | --- | --- | --- | --- | --- |
|  |  | Unadjusted |  |  | Adjusted^ |  |  |  | Unadjusted |  |  | Adjusted^ |  |  |
|  | N<br>(N<sub>admitted</sub>) | OR | 95%<br>CI | p-value | OR | 95%<br>CI | p-value | N<br>(N<sub>admitted</sub>) | OR | 95%<br>CI | p-value | OR | 95%<br>CI | p-value |
| **Age group (years)** |  |  |  |  |  |  |  |  |  |  |  |  |  |  |
| 0-39 | 654 (54) | - | - | - | - | - | - | 294 (7) | - | - | - | - | - | - |
| 40-54 | 348 (38) | 1.36 | 0.87,<br>2.10 | 0.2 | 1.61 | 1.02,<br>2.52 | 0.040 | 119 (3) | 1.06 | 0.23,<br>3.89 | >0.9 | 0.96 | 0.20,<br>3.70 | >0.9 |
| 55+ | 167 (32) | 2.63 | 1.62,<br>4.22 | <0.001 | 3.52 | 2.02,<br>6.05 | <0.001 | 59 (8) | 6.43 | 2.22,<br>19.1 | <0.001 | 5.08 | 1.32,<br>18.8 | 0.015 |
| **Gender** |  |  |  |  |  |  |  |  |  |  |  |  |  |  |
| Male | 597 (55) | - | - | - | - | - | - | 223 (5) | - | - | - | - | - | - |
| Female | 572 (69) | 1.35 | 0.93,<br>1.97 | 0.11 | 1.38 | 0.95,<br>2.03 | 0.10 | 249 (13) | 2.40 | 0.89,<br>7.59 | 0.10 | 3.17 | 1.11,<br>10.6 | 0.041 |
| **Chronic health condition\*** |  |  |  |  |  |  |  |  |  |  |  |  |  |  |
| No | 1,018<br>(104) | - | - | - | - | - | - | 432 (12) | - | - | - | - | - | - |
| Yes | 151 (20) | 1.34 | 0.78,<br>2.20 | 0.30 | 0.93 | 0.51,<br>1.63 | 0.8 | 40 (6) | 6.18 | 2.04,<br>17.0 | <0.001 | 2.86 | 0.76,<br>10.4 | 0.11 |
| **COVID-19 vaccination status<sup>#</sup>** |  |  |  |  |  |  |  |  |  |  |  |  |  |  |
| Not vaccinated | 568 (73) | - | - | - | - | - | - | 44 (1) | - | - | - | - | - | - |
| Partially vaccinated | 198 (24) | 0.95 | 0.57, 1.54 | 0.8 | 0.73 | 0.43, 1.22 | 0.2 | 25 (2) | 3.74 | 0.34, 83.0 | 0.3 | 4.41 | 0.37, 103 | 0.3 |
| Fully vaccinated | 403 (28) | 0.51 | 0.32, 0.80 | 0.004 | 0.40 | 0.25, 0.64 | <0.001 | 403 (15) | 1.66 | 0.32, 30.4 | 0.6 | 1.36 | 0.24, 25.0 | 0.8 |

OR = odds ratio, CI = confidence interval. ^All listed variables were included in the multivariable (adjusted) model: age, gender, vaccination status, chronic health condition and admission to hospital.
\* self-reported diabetes, cardiovascular disease, HIV, cancer, or asthma.

### Global Context

We identified closely related genomes to understand the origins of the Nepal strains within the global SARS-CoV-2 context. We linked 687 genomes from countries beyond Nepal to our samples using a three single nucleotide polymorphism (SNP) and three-week threshold (see Supplementary Materials Figure S2). These linked samples were collected in 43 different countries between 1 February 2021 and 28 October 2022, shown in Table 3. India had the highest number of linked samples (n=233; 33.9%). Other countries known to be destination countries for migrant workers from Nepal were identified with a high proportion of the linked samples, including Malaysia, Bangladesh, United Arab Emirates and Qatar, shown in Table 3 (8, 9). Interestingly, the Philippines had a high number of linked samples i.e. two samples collected in Nepal were linked to several samples in the Philippines (see Supplementary Materials Figure S2).

**Table 3:** Country of origin for SARS CoV-2 genomes that are within three SNPs and sampled within three weeks of a sample from our dataset. The total number of samples meeting the defined threshold for links for each variant (Delta/Omicron), total number of samples sequenced in the country throughout the pandemic (Total sequencing capacity), the percentage of total sequencing capacity that were linked to samples in the Epidemic Intelligence study (Percent of capacity). Countries with greater than 0.1% of their total sequencing capacity linked to Nepal are highlighted in green.

| Country | Delta links | Omicron links | Total links | Total sequencing capacity | Percent of capacity |
| --- | --- | --- | --- | --- | --- |
| India | 145 | 88 | 233 | 221127 | 1.05E-01 |
| Philippines | 95 |  | 95 | 28561 | 3.33E-01 |
| Germany | 41 | 38 | 79 | 1592069 | 4.96E-03 |
| Italy | 39 |  | 39 | 179735 | 2.17E-02 |
| Australia |  | 37 | 37 | 223775 | 1.65E-02 |
| USA | 11 | 24 | 35 | 4999958 | 7.00E-04 |
| Japan | 20 | 5 | 25 | 675109 | 3.70E-03 |
| Turkey |  | 20 | 20 | 73371 | 2.73E-02 |
| Indonesia |  | 19 | 19 | 48843 | 3.89E-02 |
| England | 14 | 2 | 16 | 2400611 | 6.66E-04 |
| United Arab Emirates | 15 |  | 15 | 6612 | 2.27E-01 |
| Bangladesh |  | 14 | 14 | 7287 | 1.92E-01 |
| Denmark | 5 | 8 | 13 | 659323 | 1.97E-03 |
| Malaysia |  | 8 | 8 | 41513 | 1.93E-02 |
| Mozambique | 7 |  | 7 | 1733 | 4.04E-01 |
| New Zealand |  | 7 | 7 | 15337 | 4.56E-02 |
| Canada | 6 |  | 6 | 625645 | 9.59E-04 |
| Singapore | 3 |  | 3 | 49319 | 6.08E-03 |
| Belgium | 3 |  | 3 | 166123 | 1.81E-03 |
| Qatar | 2 |  | 2 | 5534 | 3.61E-02 |
| Iceland | 2 |  | 2 | 40161 | 4.98E-03 |
| South Africa | 2 |  | 2 | 50334 | 3.97E-03 |
| Sweden |  | 2 | 2 | 258552 | 7.74E-04 |
| South Korea | 1 |  | 1 | 17001 | 5.88E-03 |
| Czech Republic |  | 1 | 1 | 36082 | 2.77E-03 |
| Finland | 1 |  | 1 | 49407 | 2.02E-03 |
| Russia | 1 |  | 1 | 80537 | 1.24E-03 |
| Brazil | 1 |  | 1 | 239395 | 4.18E-04 |
| **Total** | **414** | **273** | **687** |  |  |

**Figure 2:**
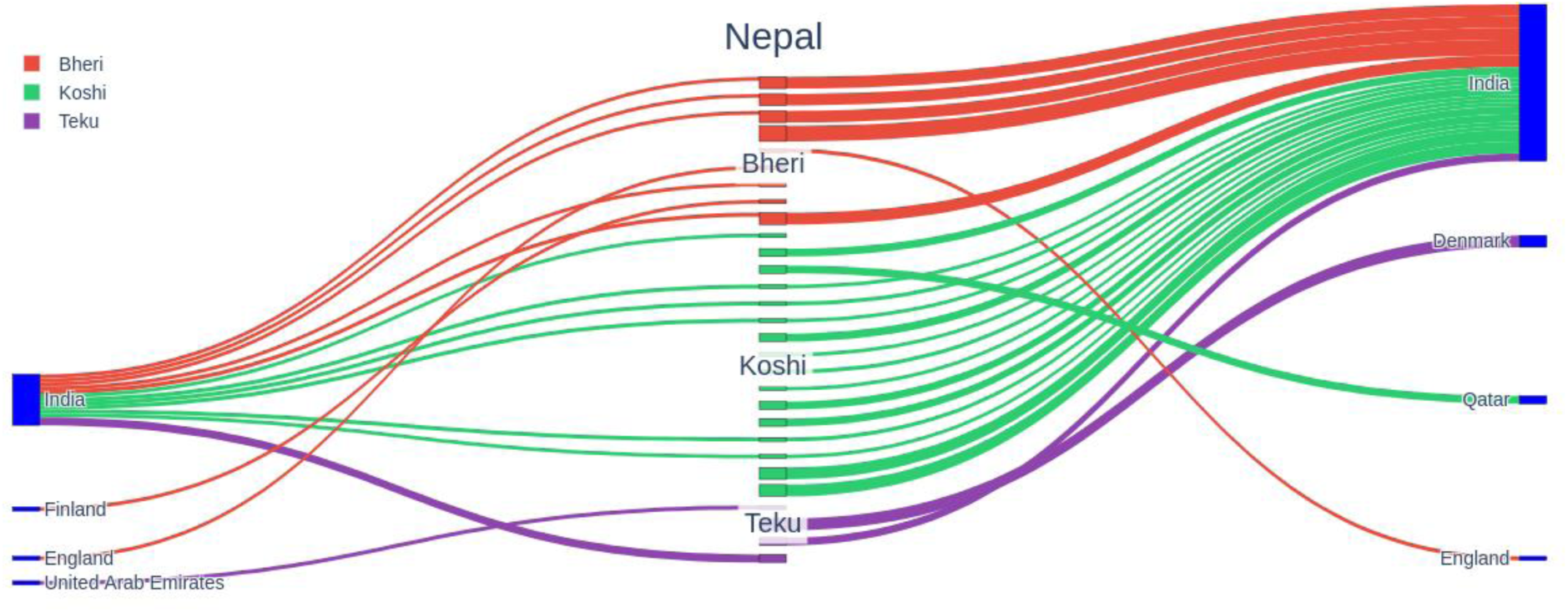
Sankey plot of samples with ‘strong’ links i.e. Nepali samples linked to a single location in the global datasets. Linked samples are defined as samples collected outside Nepal that are within three SNPs and were collected within three weeks of a sample from our study. The middle column shows individual samples collected in Nepal coloured by collection site. The left column shows countries with collection dates before the matching sample. The right column shows linked countries with collection dates on or after the collection date of the linked Nepali sample.

Recent travel history was only reported by 31 (4.5%) participants with associated genomic data in our study. Of the participants that reported recent travel history half (n=15/31; 48.3%) were linked to samples in the global dataset using the three-SNP and three-week threshold. The country of origin of most of the linked samples was Nepal itself (n=10/15; 66.6%), suggesting local transmission. Two participants reporting recent travel to India were linked to samples from India. This suggests that they may have acquired the infection while traveling in India, although the linked samples in these two cases were collected both before and after the collection date of the sample in Nepal, making it impossible to draw conclusions regarding direction of transmission.

### Phylogeography of Delta and Omicron

Asymmetric “migration” models, used in a phylogeographic framework, attempt to reconstruct directionality of transitions or exchange events between geographic locations (11). We conducted a phylogeographic analysis between India and Nepal with Nepali samples stratified by Nepali collection site: Bheri-Western Nepal, Teku-Central Nepal or Koshi-Eastern Nepal (Figure 6).

The pattern of SARS CoV-2 transmission between India and Nepal changed substantially between the two major variant waves. The emergence of Delta variant in India was followed by many introductions into Nepal. There were frequent importation events of Delta variants from India into Nepal, with relatively few domestic strain migrations within Nepal occurring predominantly from Teku (Kathmandu; the capital city) to neighbouring Nepal sites, and only rare transmission events out of Bheri, in the West of Nepal, into the other Nepali sites (Figure 3).

**Figure 3:**
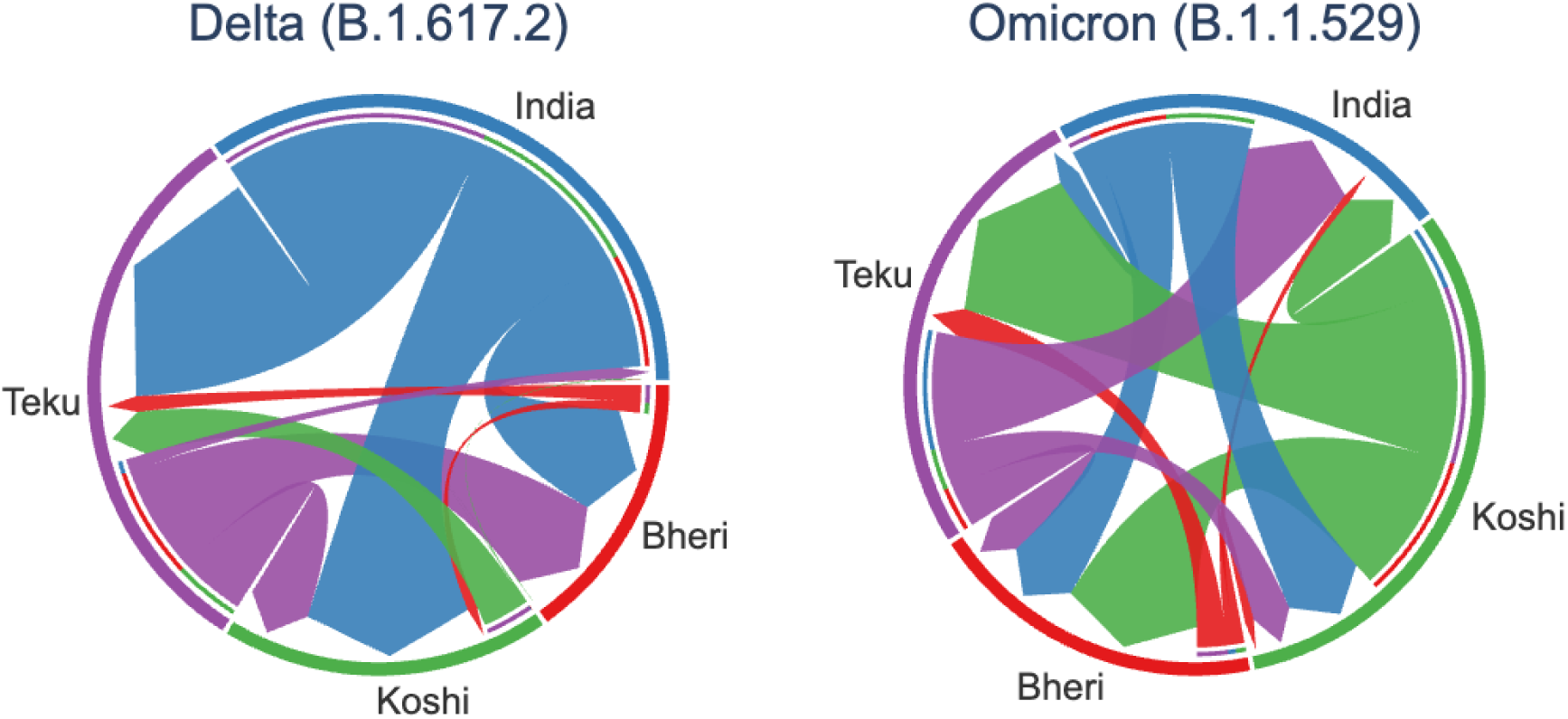
Circular migration flow plots for all sampled locations counting the number of exchange events (Markov jumps) in the phylogeographic model. The arrow heads indicate the direction of exchange. Outer circle of the plot represents the source location, and the inner circle is the destination.

During the Omicron wave, post March 2022, however, the dynamics shifted to show evidence of transmission occurring consistently between India and all Nepali sites (Figure 3). This was after the end of the major national lockdown restrictions in Nepal when internal and external travel was less inhibited (Figure 1). During the Omicron wave Koshi (Eastern Nepal) appeared to account for the largest number of exports into neighbouring sites of Nepal.

### Prevalence of self-reported long COVID

Defining long COVID with the broadest definition, and including any participants who reported initial COVID symptoms lasting over 12 weeks and a symptom specified in the questionnaire at the time of interview, resulted in a high prevalence of participants being classified as long COVID cases at both the 3- (n=871/1992; 43.7%) and 6-month (n=167/1970; 8.5%) follow up (see Supplementary Materials Table S2).

At both time points, the prevalence of post-COVID symptoms was higher for participants infected with the Delta variant: 55.0% (n=638/1161) at 3 months and 9.7% (n=112/1152) at 6 months compared to 22.5% (n=112/556) and 6.4% (n=35/546) at 3 and 6 months, respectively, for Omicron. The high prevalence of post-COVID symptoms was principally due to a high proportion of participants reporting weakness/tiredness at 3 months (n=718/1992, 36.0%) and 6 months (n=447/1970, 22.7%) (see Figure 4A). This phenomenon continued in the 12-month questionnaire, with 19.7% of participants (n=114/580) reporting weakness/tiredness.

**Figure 4:**
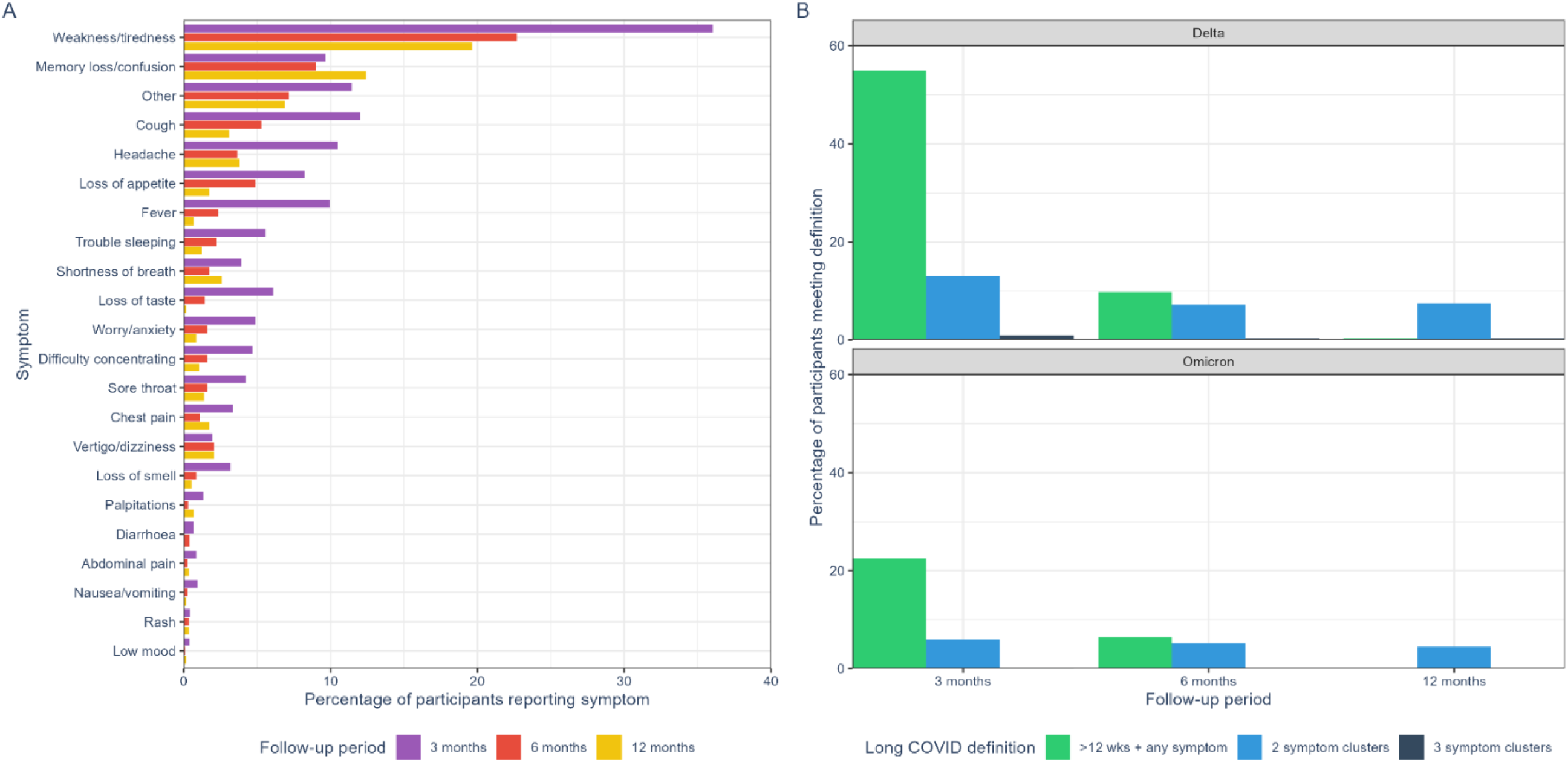
A) Percentage of participants reporting each long COVID symptom in the 3-month (N=1992), 6-month (N=1970) and 12- month (N=580) follow-up questionnaires. Participants for the 12-month questionnaire were selected based on the duration of symptoms from their initial SARS-CoV-2 infection reported in the 3-month questionnaire, see Methods. B) Percentage of participants who met each self-reported long COVID definition at 3-, 6-and 12-month follow up, by variant (data provided in Supplementary Materials Table S2). Long COVID definitions: >12 wks + any symptom = initial COVID-19 symptoms lasted >12 weeks and reported any symptom specified in Longitudinal Population Study questionnaire; 2 symptom clusters = reported at least two of the symptom clusters in the WHO Delphi definition; 3 symptom clusters = reported all three symptom clusters in the WHO Delphi definition.

Using the symptoms included in the WHO Delphi consensus definition and classifying participants who reported at least two of the symptom clusters in the WHO Delphi definition as long COVID cases resulted in 13.1% (n=152/1161) and 7.1% (n=82/1152) of participants infected with Delta being classified as reporting long COVID at 3 and 6 months, respectively. Using this definition, 5.9% (n=33/556) and 5.1% (n=28/546) of those infected with Omicron were classified as having long COVID at 3 and 6 months, respectively (see Figure 4B). This definition was used in subsequent analyses to investigate risk factors for post-covid sequelae.

We also analysed the data to identify participants reporting all three symptom clusters in this definition of long COVID cases as a proxy measure for more severe cases. Fourteen participants reported experiencing all three of the symptom clusters of long COVID at 3 months (n=14/1992; 0.7%), ten participants infected with the Delta variant (n=10/1161; 0.9%) and 1 with Omicron variant (n=1/556; 0.2%).

Including those who reported at least two symptom clusters from the WHO Delphi definition, there was considerable geographic variation in the prevalence of long COVID at 3 months and 6 months for those infected with Delta, based on location of recruitment. At 3 months, 3.1% (n=10/321) of participants recruited in Bheri reported long COVID, compared to 10.5% (n=51/495) from Koshi and 26.4% (n=91/345) from Teku. At 6 months, this pattern persisted but was less pronounced: 1.6% (n=5/319) of participants from Bheri reported long COVID, compared to 7.4% (n=36/489) from Koshi and 11.9% (n=41/344) from Teku. These differences were less pronounced for those infected with Omicron at both 3 and 6 months. At 3 months, 3.1% (n=7/227) of participants recruited in Bheri reported long COVID, compared to 8.0% (n=13/163) from Koshi and 7.8% (n=13/166) from Teku. At 6 months, 3.2% (n=7/222) of participants from Bheri reported long COVID, compared to 9.3% (n=15/161) from Koshi and 3.7% (n=6/163) from Teku.

The prevalence of long COVID at 12 months was higher for those infected with Delta compared to Omicron across all long COVID definitions (see Figure 4B), and higher among those who reported their initial COVID-19 symptoms lasted more than 12 weeks (37/289; 12.8%), compared to those who reported their symptoms lasted less than 4 weeks (11/291; 3.8%) (see Supplementary Materials Table S3).

In the 3-month follow-up questionnaire, 257 participants (of 1992 who responded; 12.9%) reported they had discussed symptoms that lasted more than 4 weeks with a doctor or nurse, 227 (11.4%) had visited a pharmacist and 249 (12.5%) had consulted another health professional.

There were 81 participants (4.9%) who reported they had developed a new condition since their COVID-19 diagnosis, of whom 15 reported they were told by a doctor that this condition developed because of COVID-19.

Some participants self-reported additional SARS-CoV-2 infections during the follow-up period, with 83 participants (4.2%) re-infected before completing the 3-month questionnaire and 41 (2.1%) before the 6-month questionnaire. The timing of follow-up questionnaire completion was not precise (details given in Supplementary Materials).

### Long COVID phylogenetics

We visualised samples from those meeting our long COVID definition on a phylogenetic tree. There were no signals of long COVID population structure within the Delta or Omicron trees, i.e. long COVID cases appeared to be randomly distributed within the two major SARS-CoV-2 lineages (see Figure 5). However, the portion of long COVID cases in each variant was not equal. To further investigate this variant discrepancy, factors associated with long COVID were statistically assessed.

**Figure 5:**
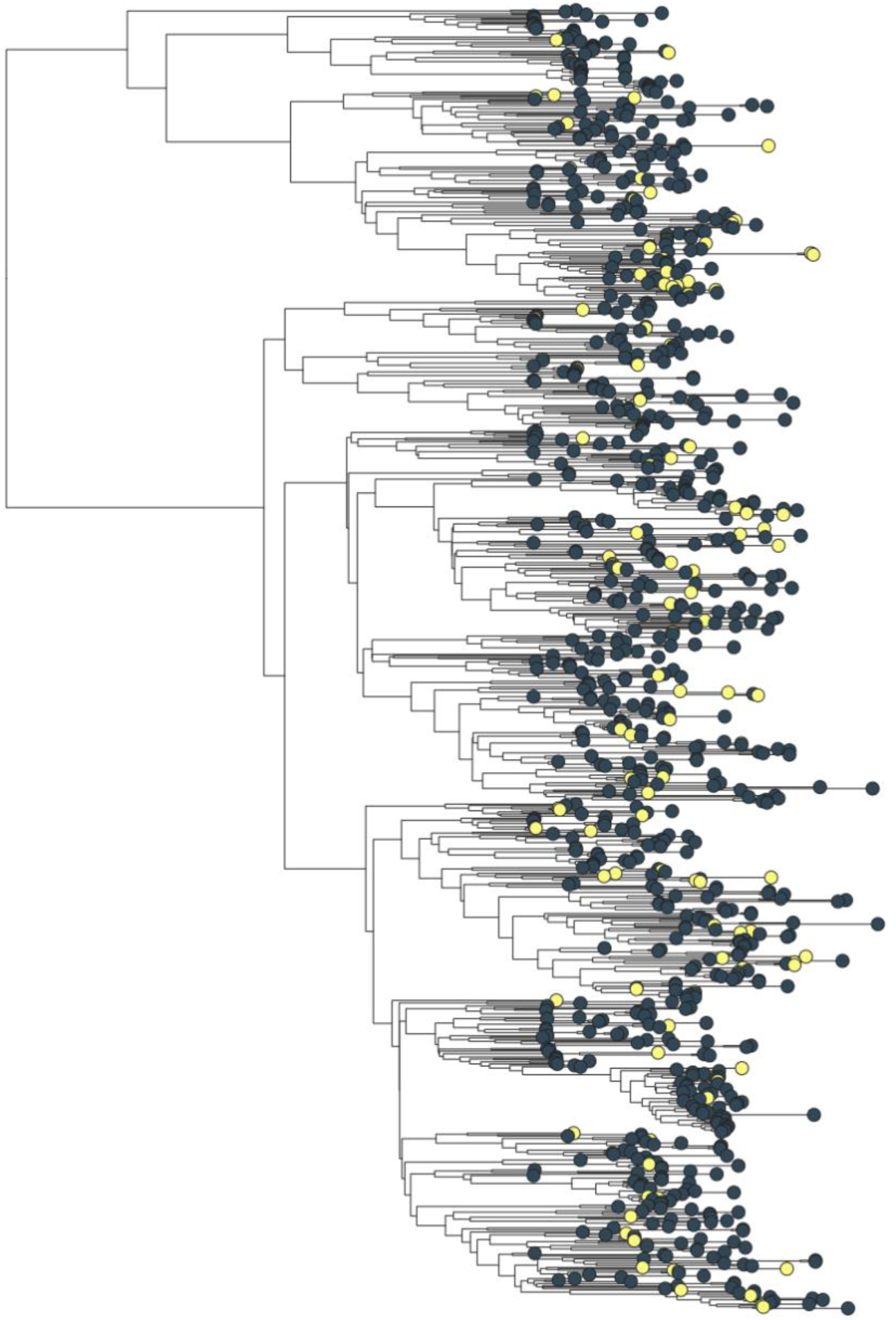
Time-scaled maximum clade credibility tree of Delta samples showing the distribution of long COVID cases in the phylogeny. Samples from patients meeting the long COVID definition (reporting at least two of the symptom clusters in the WHO Delphi definition in the 3-month questionnaire) are highlighted in yellow.

**Figure 6:**
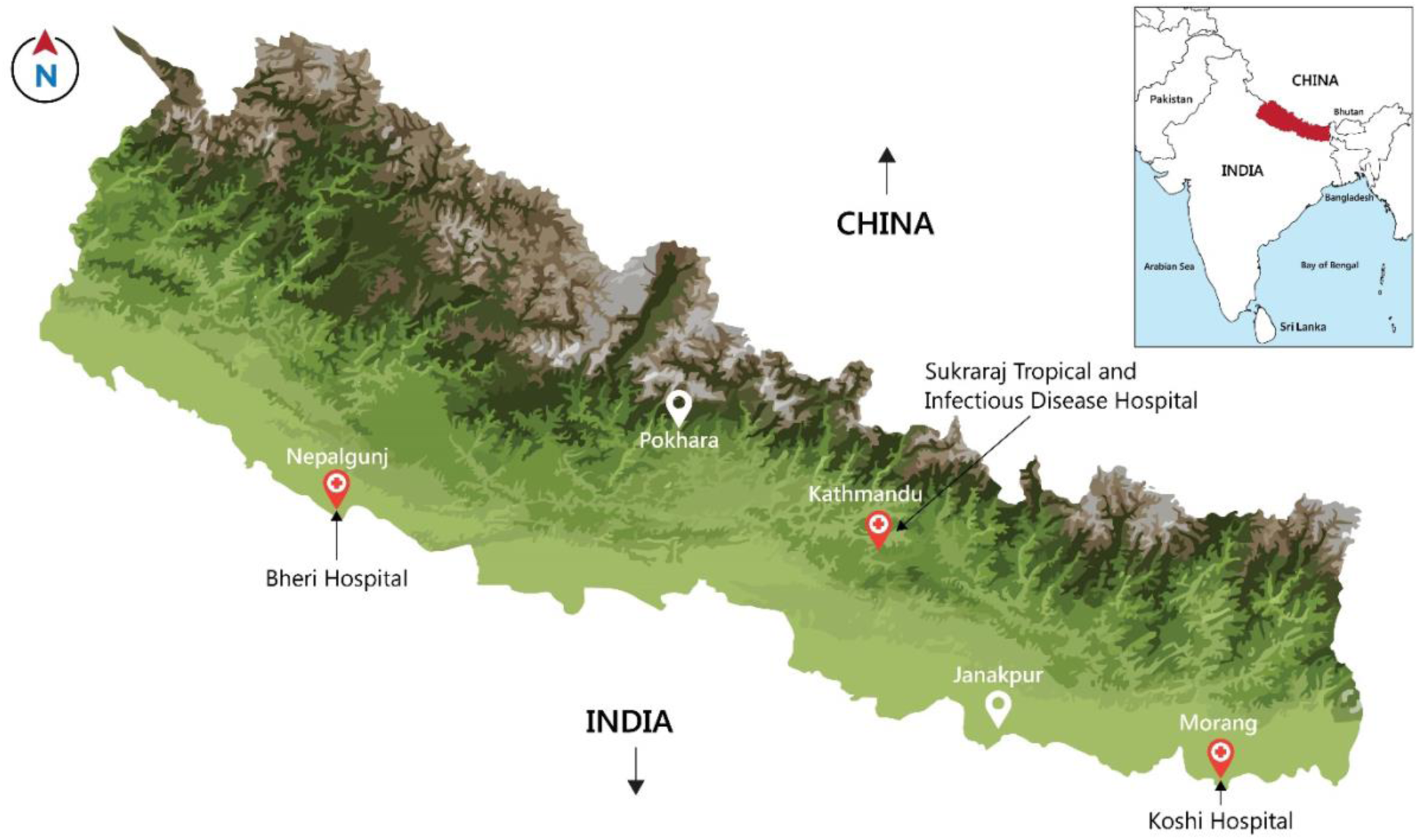
Map of Nepal showing Epidemic Intelligence study recruitment hospital locations All patients diagnosed at the hospital with COVID-19 by real time polymerase chain reaction (RT-PCR) were eligible. Eligible participants were contacted by study trained community health workers who explained the study and invited them to participate. Potential participants were provided with a participant information sheet and given time to consider participation. Those agreeing to participate provided written informed consent. If the COVID-19 patient was under 18 years of age or unable to provide consent due to a medical condition (confused or unconscious), the legal next of kin was approached for consent. Participants between 12 and 18 years of age were asked to provide assent, in addition to the written informed consent of the legal parent or guardian. COVID-19 patients diagnosed in the three months prior to the start of recruitment were also eligible to participate if the hospital had a stored sample and were approached for consent to participate.

### Factors associated with long COVID

Long COVID was more common among those infected for the first time with the Delta variant (n=148/1137; 13.0%) than for those infected with the Omicron variant (n=24/464; 5.2%). This was true across all age groups (see Table 4). For those infected with Delta, we found that female gender (adjusted OR=1.63 [95% CI 1.15, 2.34; P=0.007), having a chronic health condition (adjusted OR=1.79 [95% CI 1.09, 2.90]; P=0.019) and being fully vaccinated at the time of infection (adjusted OR=1.87 [95% CI 1.25, 2.81]; P=0.003) were associated with increased risk of self-reported long COVID at 3 months (see Table 4). Notably, among those infected with the Delta variant, the prevalence of long COVID at 3 months was higher among those who were fully vaccinated (n=69/393;17.6%) at the time of their initial SARS-CoV-2 infection compared to those who were not vaccinated (n=52/552; 9.4%) (adjusted OR=1.87 [95% CI 1.25, 2.81]; P=0.003).

**Table 4:** Factors associated with self-reported long COVID at 3 months, among those infected with Delta and Omicron who did not report a previous SARS-CoV-2 infection.

|  | Delta (N=1,137) |  |  |  |  |  |  | Omicron (N=464) |  |  |  |  |  |  |
| --- | --- | --- | --- | --- | --- | --- | --- | --- | --- | --- | --- | --- | --- | --- |
|  |  | Unadjusted |  |  | Adjusted^ |  |  |  | Unadjusted |  |  | Adjusted^ |  |  |
|  | N<br>(N<sub>LC</sub>) | OR | 95%<br>CI | p-value | OR | 95%<br>CI | p-value | N<br>(N<sub>LC</sub>) | OR | 95%<br>CI | p-value | OR | 95%<br>CI | p-value |
| **Age group (years)** |  |  |  |  |  |  |  |  |  |  |  |  |  |  |
| 0-39 | 630 (67) | - | - | - | - | - | - | 290 (8) | - | - | - | - | - | - |
| 40-54 | 343 (49) | 1.40 | 0.94,<br>2.07 | 0.094 | 1.12 | 0.74,<br>1.70 | 0.6 | 118 (13) | 4.36 | 1.79,<br>11.3 | 0.001 | 4.84 | 1.91,<br>13.0 | 0.001 |
| 55+ | 164 (32) | 2.04 | 1.27,<br>3.21 | 0.003 | 1.33 | 0.77,<br>2.27 | 0.3 | 56 (3) | 2.00 | 0.43,<br>7.15 | 0.3 | 2.13 | 0.41,<br>8.60 | 0.3 |
| **Gender** |  |  |  |  |  |  |  |  |  |  |  |  |  |  |
| Male | 580 (61) | - | - | - | - | - | - | 218 (8) | - | - | - | - | - | - |
| Female | 557 (87) | 1.57 | 1.11,<br>2.24 | 0.011 | 1.63 | 1.15,<br>2.34 | 0.007 | 246 (16) | 1.83 | 0.79,<br>4.58 | 0.2 | 2.00 | 0.84,<br>5.14 | 0.13 |
| **Chronic health condition\*** |  |  |  |  |  |  |  |  |  |  |  |  |  |  |
| No | 987 (114) | - | - | - | - | - | - | 425 (22) | - | - | - | - | - | - |
| Yes | 150 (34) | 2.24 | 1.45,<br>3.42 | <0.00<br>1 | 1.79 | 1.09,<br>2.90 | 0.019 | 39 (2) | 1.60 | 0.37,<br>4.94 | 0.5 | 1.25 | 0.26,<br>4.49 | 0.8 |
| **COVID-19 vaccination status<sup>#</sup>** |  |  |  |  |  |  |  |  |  |  |  |  |  |  |
| Not vaccinated | 552 (52) | - | - | - | - | - | - | 43 (3) | - | - | - | - | - | - |
| Partially vaccinated | 192 (27) | 1.57 | 0.95,<br>2.57 | 0.074 | 1.38 | 0.82,<br>2.30 | 0.2 | 25 (1) | 0.56 | 0.03,<br>4.62 | 0.6 | 0.48 | 0.02,<br>4.23 | 0.5 |
| Fully vaccinated | 393 (69) | 2.00 | 1.39,<br>3.02 | <0.00<br>1 | 1.87 | 1.25,<br>2.81 | 0.003 | 396 (20) | 0.71 | 0.23,<br>3.10 | 0.6 | 0.49 | 0.15,<br>2.21 | 0.3 |
| **Admitted to hospital for initial COVID-19 infection** |  |  |  |  |  |  |  |  |  |  |  |  |  |  |
| No | 1,019<br>(129) | - | - | - | - | - | - |  |  |  |  |  |  |  |
| Yes | 118 (19) | 1.32 | 0.76,<br>2.19 | 0.3 | 1.32 | 0.75,<br>2.23 | 0.3 |  |  |  |  |  |  |  |

OR = odds ratio, CI = confidence interval, LC = Long COVID. ^All variables were included in the multivariable (adjusted) model. Admission to hospital was not included in the Omicron model, as there were no participants admitted to hospital for their initial COVID-19 infection who reported long COVID.
\* self-reported diabetes, cardiovascular disease, HIV, cancer, or asthma.

Among those infected with the Omicron variant, age between 40-54 was the only factor associated with self-reported long COVID at 3 months (adjusted OR=4.84 [95% CI 1.91, 13.0], P=0.001), while age over 55 years was not a risk factor (adjusted OR=2.13 [95% CI 0.41, 8.60], P=0.3)(see Table 4).

The variables associated with long COVID did not differ in the sensitivity analyses including participants who reported a previous SARS-CoV-2 infection prior to enrolment in the study or excluding those who reported SARS-CoV-2 re-infections during the 3-month follow-up period, apart from age over 55 years for Omicron (see Supplementary Materials Tables S5-S6).

## Discussion

Our study shows that, throughout the COVID-19 pandemic, SARS-CoV-2 variants were repeatedly introduced into Nepal, circulated locally and were exported again, despite quarantine measures and lockdowns. We detected forty-five distinct sub-lineages but found no evidence that entirely new variants arose *de novo* within the country during the study. We found that Nepal samples were commonly linked to samples collected in India, in line with the intertwined economics and shared geography of these two countries. Most samples with strong genomic links to India were collected at sites along the Nepal-India border, emphasising the epidemiological importance of this shared porous border.

In the first phase of the Epidemic Intelligence project (June–September 2021), genomes were frequently linked to Delta strains from neighbouring India, Bangladesh and Gulf States, a source of returning economic migrants in the ‘reverse migration’ wave; domestic exchange between study sites was limited, and cross-border traffic appeared lower in the far-western Banke district (Bheri) than in central Kathmandu (Teku) or eastern Morang (Koshi). After March 2022, during the Omicron wave, the transmission patterns shifted to increased domestic exchange between the different Nepali regions and greater genomic links to SARS CoV-2 samples from multiple countries that are recipients for Nepali economic and academic migrants. This is consistent with the timing of the resumption of economic labour and academic migration from Nepal. These changing patterns illustrate how intertwined economies, geography and migration routes shaped the flow of SARS-CoV-2 variants through Nepal over time.

Of note, we linked two samples from Nepal to dozens of samples isolated in the Philippines, indicating recent transmission. The samples from the Philippines were only linked to samples from the Teku hospital site in Kathmandu, which is home to Nepal’s only international airport. The Philippines is a country without significant migration flows to/from Nepal, but which represents a ‘third country’ shared source of economic migrants to countries in the Middle East region (where several ‘strong’ links were identified). It is therefore possible that these strains were transmitted between migrants from both countries prior to repatriation during the reverse migrant flows. It is also likely this was not an isolated transmission event but an indication of a larger system of under sampled transmission networks. Future studies should focus on the role of genomics in understanding global disease transmission through economic migrant pathways.

This is especially important for countries, like Nepal, whose GDP is heavily dependent on migrant workers(8, 12).

Very few Epidemic Intelligence participants reported recent travel, and this may be due to fear of quarantine or being reported for violating quarantine regulations. During the first year of the pandemic, returning migrants, particularly those crossing the open land border from India were erratically detained in poorly equipped detention centres which lacked basic facilities and catering. Fear of the virus also led to stigma and ostracism of returning migrants by local communities (13). This led many people to avoid official channels and hide any history of migration. Spatial analysis of COVID-19 cases in the early phase of the pandemic during the first lockdown from 24 March-July 2020 showed cases spreading outwards from border crossing points with India (14). Although efforts were made to reassure participants that data was confidential and individual data would not be shared with authorities, we believe that migration history is likely to have been under-reported by our participants and our ability to analyse the links between migration and strain introduction was therefore limited. Despite this, we were able to analyse patterns of transmission from both domestic and international sources at a population level, finding compelling evidence of cross border transmission with India and other countries.

We investigated several definitions to measure the incidence and clinical phenotype of long COVID within the cohort at 3, 6, and 12-months following SARS-CoV-2 infection. This was due to the lack of an international standardised definition for the emerging syndrome. New data surrounding long COVID and a broad range of potential symptoms was continually emerging in the early phases of the pandemic. The diverse clinical presentation and interaction with pre-existing comorbidities has presented challenges in defining long COVID and standardising research into risk factors and clinical management. It is likely, due to the complexity of the underlying mechanisms and presentation, that multiple definitions will be needed to address different research questions and epidemiological analysis and research into standardised case definitions is ongoing globally (15).

While global estimates of long COVID incidence are difficult to standardise, our results are broadly concordant of those previously reported globally and from other studies in Nepal. A pooled analysis of 54 studies from 22 countries estimated 6.2% of those with symptomatic SARS CoV-2 infection experienced at least one of the three symptom clusters (fatigue, shortness of breath or cognitive dysfunction) at 3 months post-infection (16). Our data shows that almost half (43.7%) of the Nepali cohort experienced post COVID syndrome in some form, most commonly over 12 weeks of symptoms including persistent fatigue and exhaustion at 3 months. This estimate is comparable to another Nepalese studies, with a study of 300 patients admitted to a Kathmandu hospital in 2021 with COVID-19, which reporting that 39.3% had post COVID-19 symptoms at 3 months, defined as persistent clinical signs and symptoms that last more than 12 weeks and are not explained by another diagnosis (17), whereas a review of hospital records reported that 59% of 6,151 patients with COVID-19 had at least one symptom one month after recovery from acute COVID-19(18). While exact estimates are challenging due to variation in study design and case definitions, all available data indicate that clinically important symptoms persist beyond 12 weeks in a substantial proportion of cases, with significant implications including the impact on the health system and the economic burden. We also found a higher incidence of long COVID among those infected with Delta variant compared to the Omicron variant at both 3 months and 6 months, consistent with reports from Europe of a reduction in long COVID during the Omicron wave which was independent of vaccination status (19–21).

Our ability to measure long COVID relied on participants self-reporting symptoms, some of which are non-specific (such as “weakness/tiredness”), and we were unable to exclude other causes of these symptoms. Our study population is a convenience sample of those presenting to three hospitals for SARS-CoV-2 testing and may not provide an estimate of the prevalence of long COVID for the Nepali population. However, if even 3% of people infected with SARS CoV-2 experience long COVID, there will be significant impacts on the health system, alongside the social and financial consequences for affected individuals (22, 23). There is limited specialist clinical provision to support patients experiencing long COVID in Nepal, with referral services only available in large cities.

Of concern, a full course of vaccination during the Delta wave, although protective against hospitalisation and severe symptoms in the acute phase of the illness, appeared to increase the risk of long COVID in our cohort. Vaccination did not influence the risk of long COVID for those infected with the Omicron variant in our cohort. Another study of Nepali women also found that vaccination was more frequent in unrecovered women than among recovered women at 6 months following SARS-CoV-2 infection (24).The reasons for this are unknown, and the findings conflict with other studies globally which have shown a protective effect of prior vaccination in reducing long COVID incidence, including for both Delta and Omicron variants. Although there is variation in estimates across studies, pooled analysis suggests there is an approximately 40% [15-70%] risk reduction among individuals vaccinated prior to SARS CoV-2 infection (25). The discrepant findings may be related to the timing of vaccination availability in Nepal, which was after the first waves of infection. Immune dysregulation has been identified as an underlying mechanism driving long COVID, although the syndrome remains poorly characterised and further research on mechanisms of immunity and pathology is clearly needed. By April 2021, two-thirds of the unvaccinated Nepali population was seropositive for SARS CoV-2 antibodies (26).

Genomic epidemiology surveillance has a crucial role in monitoring for the emergence of new variants of SARS CoV-2 with increased virulence, vaccine escape mutants or changing clinical phenotypes, including changes in risk and presentation of long COVID syndromes. The COVID-19 pandemic highlighted critical pathogen genomic surveillance capacity gaps in low-and middle-income countries, which are particularly vulnerable to the emergence of novel pathogens with epidemic potential due to the high intensity human-animal interface in such countries, coupled with weak diagnostic and infection control capacities within the health systems. The emergence of long COVID also highlighted critical gaps in both health service delivery and research capacity to address post viral syndromes which have been historically neglected, with those affected facing many barriers to care, even in high income countries with advanced health systems (27, 28). Ongoing attempts to characterise and quantify the burden of long COVID are hampered by lack of funding, particularly in countries such as Nepal with already overstretched and fragile health systems struggling to respond to the dual burden of infectious diseases alongside a growing burden of non-communicable disease morbidity (29). Establishment of long-term cohorts of people affected by long COVID in low-and middle-income countries will enable robust evaluation of novel diagnostic, treatment and clinical management strategies.

Our study highlights the dynamic nature and lasting consequences of the SARS-CoV-2 pandemic in Nepal. The porous borders between India fuelled repeated viral exchange events and migration patterns appeared to shape viral flow over time, including through hidden transmission networks among third-country migrants. Long COVID is difficult to define but had a clear impact on the cohort in this study. Further research into the long-term clinical management and broader societal impacts of long COVID, particularly in low-and middle-income countries where the effects are often invisible, is urgently needed.

## Methods and Materials

### Participant recruitment

Participants were recruited from June 2021 to February 2023 at three government tertiary referral hospitals in Nepal (shown on map in Figure 6):

- Sukaraj Tropical and Infectious Disease Hospital (also known as Teku Hospital), in Kathmandu, the capital city in central Nepal
- Bheri Hospital, in Nepalgunj, Banke District, in the far western terrai (plains) region bordering India
- Koshi Zonal Hospital, in Biratnagar, Morang District, in the far eastern terrai (plains) region bordering India

Together these hospitals span Nepal with catchment populations from four Provinces: Koshi, Bagmati, Lumbini, and Karnali.

### Epidemiological data collection

Data was collected using a structured interview by study trained community health workers at four time points. All participants were interviewed at initial COVID-19 diagnosis and at 3-and 6-months following diagnosis. A subset (n=580) of participants were interviewed at 12-months post diagnosis following an extension to the study funding. These participants were selected based on the duration of symptoms from their initial SARS-CoV-2 infection reported in the 3-month questionnaire (291 participants who reported symptoms for less than four weeks and 289 with symptoms for greater than 12 weeks were selected by selecting every third eligible participant in the list). Among those approached to complete the 12-month questionnaire, 17 declined to participate and the next eligible participant was selected until the appropriate number of participants was recruited.

Field staff contacted participants by phone to obtain verbal consent to visit their home or work address to complete each questionnaire. Social distancing and mask protocols were adhered to during interviews.

Participants received 500 Nepali rupees (approximately 4 USD) upon completion of each questionnaire to compensate them for time taken to complete the interview. This figure is based on the local minimum wage and is a standard time compensation approved by the ethical review boards. Data from hard-copy questionnaires were entered into the customised, secure ONA study database (https://ona.io). Data accuracy was monitored by the study co-ordinator by randomly checking 10% of entries for accuracy against hard copy forms. Any repeated errors arising were flagged and discussed with the data entry staff to correct misunderstandings and improve consistency in data entry.

### Sample collection and processing

Participant samples were confirmed as positive by RT-PCR at the respective hospitals. Swab samples with cycle threshold (Ct) values less than or equal to 30 were transferred for sequencing. Samples were frozen in liquid nitrogen and transported to Center for Molecular Dynamics Nepal (CMDN) laboratory in Thapathali, Kathmandu for whole genome sequencing.

The samples underwent RNA extraction using 200μl abGenix viral DNA and RNA Extraction Kit, (AIT Biotech, Singapore) in an automated nucleic acid extraction system (abGenix, AITBiotech, Singapore). The extracted RNA was subsequently re-confirmed for SARS-CoV-2 positivity at CMDN using the Allplex™ RT-PCR SARS-CoV-2 Assay (Seegene Inc., Korea). The PCR was performed following the kit manual and as described in Napit et al.(30).

### SARS-CoV-2 sequencing

Samples with Ct values exceeding 30 at CMDN were excluded from further analysis. Complementary DNA (cDNA) was prepared for all the selected RNA using the Superscript cDNA synthesis kit (Thermo Fisher Scientific, USA), adhering to the manufacturer’s instructions. The cDNA was then used for amplification of entire genome of SARS-CoV-2 using ARTIC PCR protocol (Napit et.al. 2023). 98 pairs of primers were used in two pools P1 and P2 targeting ∼400bp amplicon of SARS-CoV-2 with ∼200bp overlap. Minor adjustments were made to PCR cycles according to the Ct values. The amplification was verified by checking for single distinct bands of ∼400bp size in each pool and then pooled together for each sample.

The pooled PCR products underwent a cleanup process using AMPure XP beads (Beckman Coulter, USA) followed by library preparation for sequencing with the Nextera XT library preparation kit using 1ng of purified artic products. All the products were indexed using the Nextera XT V2 index kit. The library quantification was performed in Qubit 3 (Thermo Fisher Scientific, USA) using the Qubit HS dsDNA assay kit and the library size was estimated using Agilent 1000 DNA assay kit in Agilent 2100 Bioanalyzer system (Agilent Technologies, USA). The final library was sequenced on the Illumina MiSeq system using MiSeq Reagent kit V2 (300cycles) (Illumina, USA).

### Raw sequence data processing for consensus sequence generation

The nf-core/viralrecon pipeline (v2.4.1) was used to generate SARS-CoV-2 consensus sequences from raw Illumina FASTQ data. This pipeline is a nextflow-based workflow designed for viral genome reconstruction and includes quality control, adapter trimming, and alignment to a reference genome (SARS-CoV-2 Wuhan-Hu-1 reference genome; Genbank acc: MN908947.3). It employs tools including FastQC for read quality assessment, fastp for adapter trimming, and Bowtie2 for read alignment, followed by primer sequence removal by iVar. Variant calling is performed using iVar or LoFreq, followed by consensus sequence generation and annotation.

The pipeline utilises Nextflow and Docker/Singularity. Quality control reports are generated alongside the final consensus sequences for downstream analyses.

### SARS-CoV-2 lineage assignment

Lineages were assigned using Nextclade v3.8.2 with database 2024-07-17–12-57-03Z (31). We compared this Nextclade database with a database from the time of final sample collection and found them to be concordant. We collapsed lineages to either Delta (B.1.617.2) or Omicron (B.1.1.529) using pango-collapse v0.8.2 (32), all other lineages e.g. recombinants were removed.

### Quality control

For use in the phylogenetic analysis, we further filtered the sequence dataset to only include samples with an overall Nextclade QC status of’good’ and greater than 95% coverage using augur v25.2.0 (33). Known problematic sites i.e. alignment ends, ambiguous bases, amplicon dropouts, sequencing issue, etc were masked as Ns in all sequences. UShER v0.6.3 and IQ-TREE v2.3.5 (34) were used to build maximum parsimony and maximum likelihood trees, respectively(35). The topologies of the resulting trees were visually compared to assess convergence of phylogenetic structure. A root-to-tip regression analysis was performed on the maximum likelihood tree using Clockor2 v1.9.1 to determine temporal signal and suitability for downstream phylodynamic analysis (36). Sequence data was submitted as available after Quality Control checks to the global open access GISAID database throughout the study (https://gisaid.org/hcov19-variants/).

### Global context

To contextualise the Nepal dataset within the global SARS-CoV-2 dynamics we identified closely related samples and performed an analysis of the Nepali and public samples from India.

### Inclusion of external sequence data

We compared the SARS CoV-2 genomes sampled in Nepal to the 16 million genomes publicly available in global databases (GISAID, GenBank, COG-UK and CNCB) using phylogenetic placement (37)(11). Closely related genomes are highly informative for transmission events as the shared ancestry of rapidly evolving organisms approximates transmission chains. Linked samples were identified by searching a global SARS-CoV-2 phylogeny for samples within three SNPs of the samples in our dataset.

Most samples linked to the Delta dataset were from India while most of the samples linked to the Omicron dataset were sequenced in Australia. However, the Australian-Omicron samples were not temporally linked to their Nepali counterparts. That is, most of the linked Australian samples were collected more than 50 days after the Nepal samples. There could be many transmission events within a 50-day period which would break the epidemiological links between samples. It is also possible that the large number of linked samples from Australia were due to the high sequence capacity and importation events in Australia during this time (38). To create a more closely temporally linked dataset and reduce bias introduced from countries with high sequencing capacity, we filtered the dataset so that only samples collected within three weeks of each other were included. A second more stringent filtering was then applied to identify ‘strong’ links to single countries.

### Phylodynamic analysis

Sequence data from India was downloaded from GISAID using GISAIDR v0.9.10 (39). To minimise bias from over-representation of specific sampling locations, we down sampled the Delta dataset to include the same number of samples (n=200) from India and the three Nepal sampling sites (40). The Omicron dataset was already sufficiently balanced and so all high-quality samples were included (India=98, Bheri=105, Teku=91, Koshi=110). A list of GISAID IDs used in this study is provided in the Supplementary Materials (Supplementary File 1).

Phylodynamic analysis was performed on Delta and Omicron datasets separately. In both cases a skygrid demographic model from BEAST v1.10.4 was used to generate a posterior distribution of trees that was used as the bases of the subsequent discrete trait analysis (41).The analysis was run in duplicate and confirmed to reach the same stationary distribution using Beastiary v1.8.3 (42). Prior sensitivity analysis was used to bolster choice of strict clock rate prior by comparing an informative and uninformative gamma distribution.

### Phylogeographic discrete trait analysis

The empirical distribution of trees generated in the phylodynamic analysis was used as the bases for the phylogeographic analysis. PrioriTree (https://jsigao.shinyapps.io/prioritree) (43) was used to configure a continuous-time Markov chain with an asymmetric rate-matrix to describe the evolution of geographic areas (demes) over time in BEAST v1.10.4 (44). Four equally sized demes were included in the analysis (three Nepal locations and India). The number of Markov jumps and rewards were logged for each deme. We approximated the joint posterior distribution using Markov chain Monte Carlo (MCMC), running the analysis for 100 million generations and sampling every 10,000 generations throughout the chain. We ran two independent MCMC chains to assess the convergence among replicates.

### Epidemiological analysis

The laboratory and questionnaire data were compared for discrepancies in the recorded specimen collection date (assumed to be due to the use of a stored sample). Where the difference was less than 7 days, the date in the laboratory dataset was used. Where the difference was greater than 7 days, the date that was closer to the regression line in the root-to-tip regression was used.

Participants were classified as having a chronic health condition if they self-reported diabetes, cardiovascular disease, HIV, cancer, or asthma in the initial questionnaire.

Participants were classified as having a previous SARS-CoV-2 infection if the self-reported diagnosis date was more than 28 days before the specimen collection date for the positive sample included in this study, or if they reported a previous infection and no diagnosis date was provided.

Participants were classified as having a re-infection during follow-up if the self-reported diagnosis date of a SARS-CoV-2 infection was more than 28 days after the specimen collection date for the positive sample included in this study.

Participants were classified as fully vaccinated for COVID-19 if they had received the primary course of a vaccine (1 dose of the Johnson & Johnson vaccine or 2 doses of COVISHIELD, SINOVAC, Pfizer or Sinopharm vaccines), and partially vaccinated if they had received at least one dose but not the complete primary course of a vaccine (1 dose of COVISHIELD, SINOVAC, Pfizer or Sinopharm vaccines) at the time of specimen collection.

At the time of the Epidemic Intelligence study, there was no consensus definition of long COVID (45, 46). The study aimed to understand the scope of symptoms associated with long COVID and establish a cohort for further longitudinal evaluation, and therefore used a broad symptom questionnaire developed by the Wellcome Trust Longitudinal Population Survey provided by the Wellcome Longitudinal Population Study Covid-19 Steering Group and Secretariat, April 2021 version (221574/Z/20/Z) (47). This questionnaire was designed to be sensitive, rather than specific, to gather data which informed the understanding of long COVID clinical presentation. For analysis, we compared the prevalence of self-reported long COVID in our study population using three definitions based on those proposed in the academic literature.

The three definitions used to characterise long COVID using questionnaire data on self-reported symptoms collected at 3, 6 and 12 months were:

1) Reporting that initial COVID-19 symptoms lasted more than 12 weeks (based on the National Institute for Clinical Excellence UK (NICE) definition of ‘experiencing greater than 12 weeks of symptoms’(48)) and the presence of any symptom specified in the questionnaire at the time of interview (fever, headache, skin rashes, weakness/tiredness, nausea/vomiting, abdominal pain, diarrhoea, loss of appetite, loss of taste, loss of smell, sore throat, cough, shortness of breath, chest pain, palpitations, vertigo/dizziness, worry/anxiety, low mood/not enjoying anything, trouble sleeping, memory loss/confusion, difficulty concentrating)
2) Reporting the presence of at least two symptom clusters in the questionnaire that correspond to the “common symptoms” specified in the WHO clinical case definition of post COVID-19 condition by a Delphi consensus, published in October 2021 (49):

- weakness/tiredness (listed as “fatigue” in the WHO Delphi definition) AND
- shortness of breath (listed as “shortness of breath” in the WHO Delphi definition) AND
- difficulty concentrating OR memory loss/confusion (listed as “cognitive dysfunction” in the WHO Delphi definition)

3) Reporting the presence of all three symptom clusters specified in the WHO clinical case definition of post COVID-19 condition by a Delphi consensus that correspond to the three “common symptoms” (49):

- weakness/tiredness (listed as “fatigue” in the WHO Delphi definition)
- shortness of breath (listed as “shortness of breath” in the WHO Delphi definition)
- difficulty concentrating OR memory loss/confusion (listed as “cognitive dysfunction” in the WHO Delphi definition)

Univariable and multivariable logistic regression models were created using the glm function in R to explore the factors associated with hospital admission at initial infection, and the factors associated with long COVID (defined as reporting at least two of the symptom clusters in the questionnaire that correspond to the “common symptoms” specified in the WHO Delphi definition in the 3-month questionnaire). Models were created separately for those infected with Delta and Omicron, and excluded participants who reported a previous SARS-CoV-2 infection prior to enrolment. Sensitivity analyses were conducted to include those who reported a previous SARS-CoV-2 infection prior to enrolment (see Supplementary Materials Tables S4-S6). For the long COVID outcome, further sensitivity analyses were conducted, excluding those who reported a SARS-CoV-2 infection during the 3-month follow-up period.

## Ethical approval

The study was approved by the Social Welfare Council (the government authority responsible for approving all NGO projects in Nepal), the Ministry of Health and Population, Nepal Health Research Council (the government authority for approving research involving human participants in Nepal; approval number 314/202 P) and Liverpool School of Tropical Medicine Research Ethics Committee (approval number 21-047)

## Data Availability

The genomic data underlying this article are available in GISAID at gisaid.org, and all accession numbers are provided in Supplementary Material online. All data produced in the present study are available upon reasonable request to the authors.

## Acknowledgements

We would like to thank the participants and their families, and the many healthcare staff at Koshi, Bheri and Teku hospitals who facilitated sample collection and recruitment during a time of unprecedented pressure and unique challenges for healthcare personnel. We would also like to thank the government stakeholders who supported the project and enabled the necessary bureaucracy during challenging circumstances. Many staff at Centre for Molecular Dynamics contributed to sample processing and analysis, and we would like to particularly thank Rajesh Man Rajbhandari (Research Associate), Bindiya Bajracharya (Bioinformatics Officer), Saman Man Pradhan and Ashok Chaudhary (Senior Research Associates).

## Funding

The Epidemic Intelligence project was funded by the UK Foreign, Commonwealth & Development Office, UK and the Wellcome Trust, UK with an Epidemic Preparedness - Coronavirus grant, ‘Epidemic Intelligence: Understanding how returning migrant waves drive epidemic seeding and community transmission events in the South Asian context to inform epidemic preparedness’. Grant number: 222105/Z/20/Z

### Center for Molecular Dynamics Nepal

Prajwol Manandhar ^3^, Rajindra Napit ^3^, Sameer Mani Dixit ^3^, Roji Raut ^3^, Pragun Gopal Rajbhandari ^3^, Bivisha Khadka ^3^, Anupama Gurung ^3^, Seily Shrestha ^3^, and Sapana Shakya ^3^

^3^ Center for Molecular Dynamics Nepal (CMDN), Kathmandu, Nepal

