## Supplementary Materials for "The geospatial phylodynamics of SARS-CoV-2 in Nepal and clinical phenotype of long COVID during the Delta-Omicron waves"

#### COVID-19 symptoms reported in baseline questionnaire

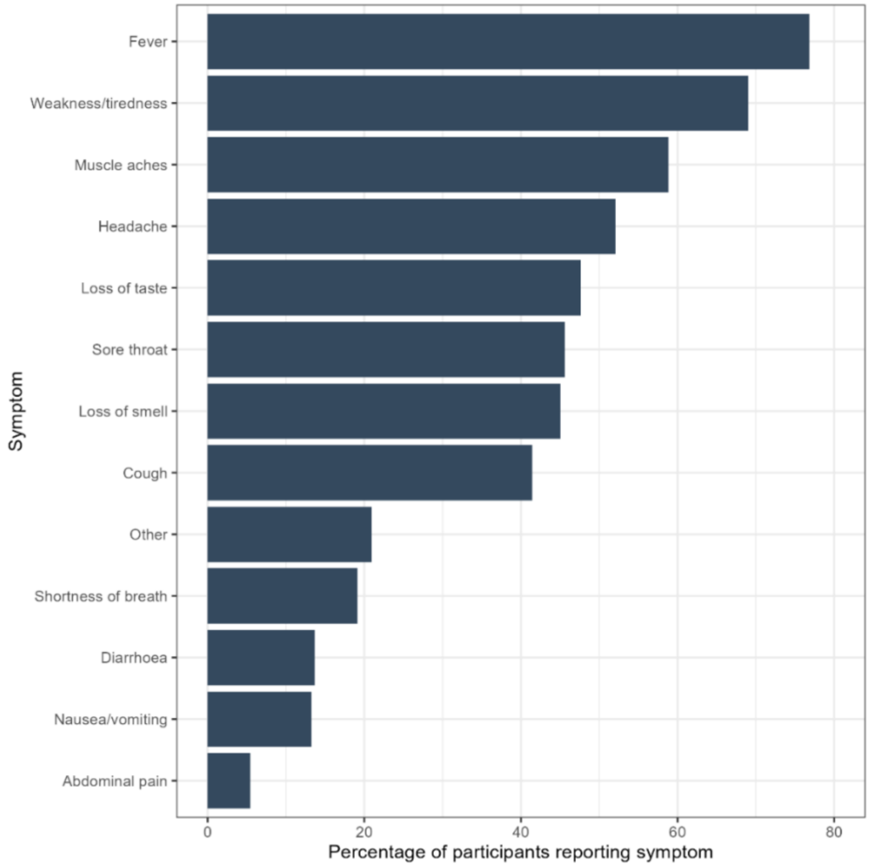

Figure S1: Percentage of participants reporting COVID-19 symptoms in the baseline questionnaire (N=2,046). Symptoms not specified in the questionnaire and captured as “other” (n=428 participants) included “runny nose” (n=203), “loss of appetite” (n=89), “sweating” (n=76), and “common cold” (n=49).

#### Study population and follow up

The time between specimen collection and completion of follow up questionnaires varied. The median interval was 161 days (5.3 months) for the 3-month questionnaire, 225 days (7.4 months) for the 6-month questionnaire and 404 days (13.3 months) for the 12-month questionnaire.

Participation and available data for each component of the study is described in Figure S2.

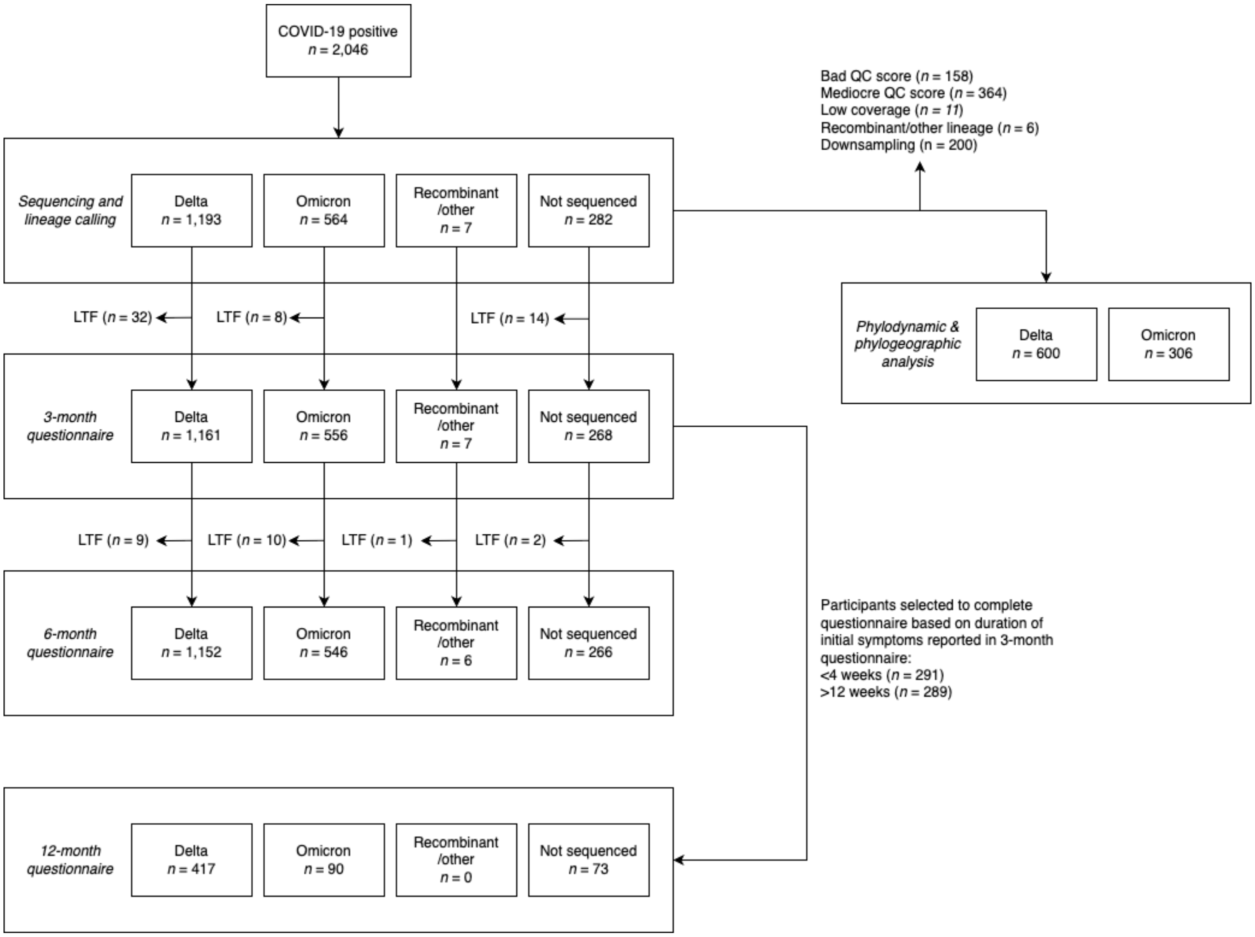

Figure S2: Participants included at each stage of the study

LFT = lost to follow-up; QC = quality control

#### Lineages identified

Table S1. Counts of Pango linages assigned to the genomes collected in this study. The lineages are in expanded lineage format where the delimiter (:) separates each alias level in the full lineage.

| Pango Lineage | Count |
|---|---|
| B.1.1.529:BA.2 | 37 |
| B.1.1.529:BA.2.10 | 35 |
| B.1.1.529:BA.2.10.1 | 1 |
| B.1.1.529:BA.2.12.1:BG.2 | 1 |
| B.1.1.529:BA.2.2 | 6 |
| B.1.1.529:BA.2.37 | 1 |
| B.1.1.529:BA.2.38 | 8 |
| B.1.1.529:BA.2.38.1 | 3 |
| B.1.1.529:BA.2.38.2 | 4 |
| B.1.1.529:BA.2.38.3 | 11 |
| B.1.1.529:BA.2.38.3:BH.1 | 4 |
| B.1.1.529:BA.2.74 | 10 |
| B.1.1.529:BA.2.75 | 26 |
| B.1.1.529:BA.2.75.1 | 24 |
| B.1.1.529:BA.2.75.1:BL.1 | 13 |
| B.1.1.529:BA.2.75.1:BL.3 | 5 |
| B.1.1.529:BA.2.75.2 | 9 |
| B.1.1.529:BA.2.75.3 | 39 |
| B.1.1.529:BA.2.75.3:BM.1.1.3 | 10 |
| B.1.1.529:BA.2.75.3:BM.2 | 1 |
| B.1.1.529:BA.2.75.3:BM.3 | 6 |
| B.1.1.529:BA.2.75.3:BM.5 | 2 |
| B.1.1.529:BA.2.75.5 | 1 |
| B.1.1.529:BA.2.75.5:BN.1 | 1 |
| B.1.1.529:BA.2.75.6 | 1 |
| B.1.1.529:BA.2.76 | 17 |
| B.1.1.529:BA.5.1 | 2 |
| B.1.1.529:BA.5.2 | 23 |
| B.1.1.529:BA.5.2.1 | 1 |
| B.1.1.529:BA.5.2.1:BF.14 | 1 |
| B.1.1.529:BA.5.2.1:BF.20 | 1 |
| B.1.1.529:BA.5.2.16 | 1 |
| B.1.1.529:BA.5.3.1:BE.4 | 1 |
| B.1.617.2 | 810 |
| B.1.617.2:AY.1 | 10 |
| B.1.617.2:AY.106 | 7 |
| B.1.617.2:AY.111 | 1 |
| B.1.617.2:AY.116 | 1 |
| B.1.617.2:AY.120 | 2 |
| B.1.617.2:AY.122 | 31 |
| B.1.617.2:AY.5 | 4 |
| B.1.617.2:AY.55 | 1 |

| | |
|---|---|
| B.1.617.2:AY.59 | 3 |
| B.1.617.2:AY.75 | 46 |
| B.1.617.2:AY.75.2 | 4 |
| **Grand Total** | **1226** |

#### Self-reported long COVID-19

Table S2: Number and percentage of participants who met each self-reported long COVID definition at 3- , 6- and 12-month follow up, by lineage (R = Recombinant/other, NS = Not sequenced, LPS = Longitudinal Population Study)

| | **3-month questionnaire** | | | | |
|---|---|---|---|---|---|
| **Long COVID definition** | **Delta (N=1,161)** | **Omicron (N=556)** | **R (N=7)** | **NS (N=268)** | **Total (N=1992)** |
| Initial COVID-19 symptoms lasted >12 weeks and reported any symptom specified in LPS questionnaire | 638 (55%) | 125 (22%) | 0 (0%) | 108 (40%) | 871 (43.7%) |
| Reported at least two of the symptom clusters in the WHO Delphi definition | 152 (13%) | 33 (5.9%) | 0 (0%) | 26 (9.7%) | 211 (10.6%) |
| Reported all three symptom clusters in the WHO Delphi definition | 10 (0.9%) | 1 (0.2%) | 0 (0%) | 3 (1.1%) | 14 (0.7%) |

| | **6-month questionnaire** | | | | |
|---|---|---|---|---|---|
| **Long COVID definition** | **Delta (N=1,152)** | **Omicron (N=546)** | **R (N=6)** | **NS (N=266)** | **Total (N=1970)** |
| Initial COVID-19 symptoms lasted >12 weeks and reported any symptom specified in LPS questionnaire | 112 (9.7%) | 35 (6.4%) | 0 (0%) | 20 (7.5%) | 167 (8.5%) |
| Reported at least two of the symptom clusters in the WHO Delphi definition | 82 (7.1%) | 28 (5.1%) | 0 (0%) | 18 (6.8%) | 128 (6.5%) |
| Reported all three symptom clusters in the WHO Delphi definition | 4 (0.3%) | 0 (0%) | 0 (0%) | 0 (0%) | 4 (0.2%) |

| | **12-month questionnaire** | | | |
|---|---|---|---|---|
| **Long COVID definition** | **Delta (N=417)** | **Omicron (N=90)** | **NS (N=73)** | **Total (N=580)** |
| Initial COVID-19 symptoms lasted >12 weeks and reported any symptom specified in LPS questionnaire | 1 (0.2%) | 0 (0%) | 0 (0%) | 1 (0.2%) |
| Reported at least two of the symptom clusters in the WHO Delphi definition | 31 (7.4%) | 4 (4.4%) | 13 (18%) | 48 (8.3%) |
| Reported all three symptom clusters in the WHO Delphi definition | 1 (0.2%) | 0 (0%) | 1 (1.4%) | 2 (0.3%) |

Table S3: Number and percentage of participants who met each self-reported long COVID definition at 12-month follow up, by duration of initial COVID-19 symptoms reported in the 3-month questionnaire (LPS = Longitudinal Population Study).

| **Long COVID definition** | **Less than 4 weeks (N=291)** | **More than 12 weeks (N=289)** |
|---|---|---|
| Initial COVID-19 symptoms lasted >12 weeks and reported presence of any symptom specified in LPS questionnaire | 1 (0.3%) | 0 (0%) |
| Reported at least two of the symptom clusters in the WHO Delphi definition | 11 (3.8%) | 37 (13%) |
| Reported all three symptom clusters in the WHO Delphi definition | 1 (0.3%) | 1 (0.3%) |

### Global context

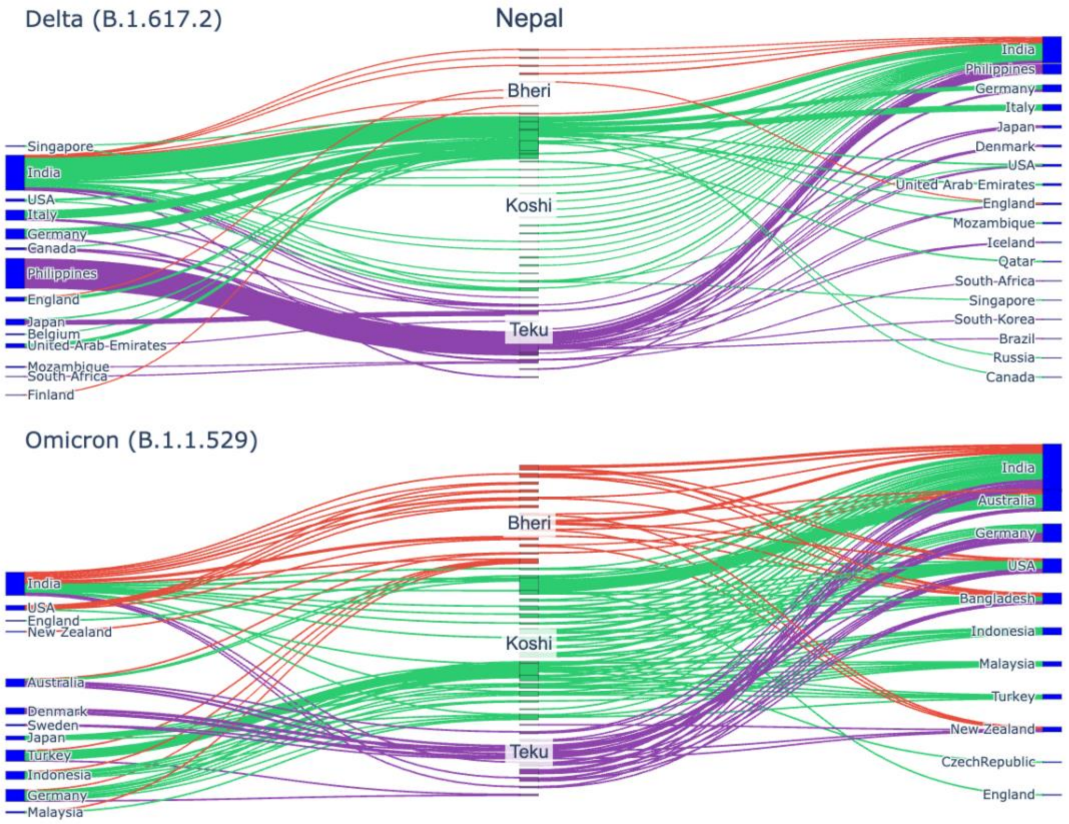

Figure S3: Sankey plot of the locations of linked samples for Delta lineage strains (top) and Omicron lineage strains (bottom) showing the changing patterns of international transmission between the waves. Linked samples are defined as samples collected outside Nepal that are within three SNPs and were collected within three weeks of a sample from the Epidemic Intelligence study. The middle column shows individual samples collected in Nepal coloured by collection site. The left column shows countries with collection dates before the matching sample. The right column shows countries with collection dates on or after the linked Nepali sample.

#### Sensitivity analyses

##### Factors associated with hospital admission

Table S4: Factors associated with hospital admission, for those infected with Delta and Omicron, including those who reported a previous SARS-CoV-2 infection

| | Delta (N=1,193) | | | | | | | Omicron (N=564) | | | | | | |
|---|---|---|---|---|---|---|---|---|---|---|---|---|---|---|
| | | Unadjusted | | | Adjusted^ | | | | Unadjusted | | | Adjusted^ | | |
| | N ($N_{admitted}$) | OR | 95% CI | p-value | OR | 95% CI | p-value | N ($N_{admitted}$) | OR | 95% CI | p-value | OR | 95% CI | p-value |
| **Age group (years)** | | | | | | | | | | | | | | |
| 0-39 | 672 (56) | - | - | - | - | - | - | 347 (8) | - | - | - | - | - | - |
| 40-54 | 352 (39) | 1.37 | 0.89, 2.10 | 0.2 | 1.61 | 1.03, 2.51 | 0.036 | 147 (3) | 0.88 | 0.19, 3.10 | 0.9 | 0.85 | 0.18, 3.12 | 0.8 |
| 55+ | 169 (33) | 2.67 | 1.66, 4.24 | <0.001 | 3.60 | 2.09, 6.14 | <0.001 | 70 (10) | 7.06 | 2.68, 19.2 | <0.001 | 5.76 | 1.70, 19.0 | 0.004 |
| **Gender** | | | | | | | | | | | | | | |
| Female | 582 (72) | - | - | - | - | - | - | 298 (15) | - | - | - | - | - | - |
| Male | 611 (56) | 0.71 | 0.49, 1.03 | 0.075 | 0.70 | 0.48, 1.02 | 0.061 | 266 (6) | 0.44 | 0.15, 1.09 | 0.090 | 0.34 | 0.11, 0.90 | 0.038 |
| **Chronic health condition*** | | | | | | | | | | | | | | |
| Male | 611 (56) | - | - | - | - | - | - | 266 (6) | - | - | - | - | - | - |
| Female | 582 (72) | 1.40 | 0.97, 2.03 | 0.075 | 1.43 | 0.98, 2.09 | 0.061 | 298 (15) | 2.30 | 0.92, 6.52 | 0.090 | 2.92 | 1.11, 8.75 | 0.038 |
| **COVID-19 vaccination status#** | | | | | | | | | | | | | | |
| Not vaccinated | 572 (73) | - | - | - | - | - | - | 49 (2) | - | - | - | - | - | - |
| Partially vaccinated | 201 (24) | 0.93 | 0.56, 1.50 | 0.8 | 0.73 | 0.43, 1.20 | 0.2 | 26 (2) | 1.96 | 0.22, 17.2 | 0.5 | 2.81 | 0.29, 27.2 | 0.3 |

| Fully vaccinated | 420 (31) | 0.54 | 0.35, 0.84 | 0.007 | 0.43 | 0.27, 0.68 | <0.001 | 489 (17) | 0.85 | 0.23, 5.44 | 0.8 | 0.79 | 0.20, 5.35 | 0.8 |
|---|---|---|---|---|---|---|---|---|---|---|---|---|---|---|

OR = odds ratio, CI = confidence interval. ^All variables were included in the multivariable (adjusted) model.

* self-reported diabetes, cardiovascular disease, HIV, cancer, or asthma.

### partially vaccinated = received some but not all the primary course of a vaccine at time of specimen collection; fully vaccinated = received the primary course of a vaccine at time of specimen collection

#### Factors associated with Long COVID

Table S5: Factors associated with Long COVID at 3 months, among those infected with Delta and Omicron, including those who reported a previous SARS-CoV-2 infection

| | **Delta (N=1,161)** | | | | | | | **Omicron (N=556)** | | | | | | |
|---|---|---|---|---|---|---|---|---|---|---|---|---|---|---|
| | | **Unadjusted** | | | **Adjusted^** | | | | **Unadjusted** | | | **Adjusted^** | | |
| | **N ($N_{LC}$)** | **OR** | **95% CI** | **p-value** | **OR** | **95% CI** | **p-value** | **N ($N_{LC}$)** | **OR** | **95% CI** | **p-value** | **OR** | **95% CI** | **p-value** |
| **Age group (years)** | | | | | | | | | | | | | | |
| 0-39 | 648 (68) | - | - | - | - | - | - | 343 (10) | - | - | - | - | - | - |
| 40-54 | 347 (51) | 1.47 | 0.99, 2.16 | 0.052 | 1.18 | 0.78, 1.77 | 0.4 | 146 (14) | 3.53 | 1.54, 8.38 | 0.003 | 3.69 | 1.58, 8.95 | 0.003 |
| 55+ | 166 (33) | 2.12 | 1.33, 3.32 | 0.001 | 1.39 | 0.81, 2.35 | 0.2 | 67 (9) | 5.17 | 1.97, 13.4 | <0.001 | 5.18 | 1.77, 14.8 | 0.002 |
| **Gender** | | | | | | | | | | | | | | |
| Male | 594 (62) | - | - | - | - | - | - | 261 (12) | - | - | - | - | - | - |
| Female | 567 (90) | 1.62 | 1.15, 2.30 | 0.006 | 1.68 | 1.18, 2.40 | 0.004 | 295 (21) | 1.59 | 0.78, 3.40 | 0.2 | 1.82 | 0.87, 3.96 | 0.12 |
| **Chronic health condition*** | | | | | | | | | | | | | | |
| No | 1,009 (117) | - | - | - | - | - | - | 507 (27) | - | - | - | - | - | - |

| | | | | | | | | | | | | | | |
|---|---|---|---|---|---|---|---|---|---|---|---|---|---|---|
| Yes | 152 (35) | 2.28 | 1.48, 3.46 | <0.001 | 1.79 | 1.09, 2.88 | 0.018 | 49 (6) | 2.48 | 0.89, 5.98 | 0.058 | 1.24 | 0.40, 3.37 | 0.7 |
| **COVID-19 vaccination status#** | | | | | | | | | | | | | | |
| Not vaccinated | 556 (52) | - | - | - | - | - | - | 48 (3) | - | - | - | - | - | - |
| Partially vaccinated | 195 (27) | 1.56 | 0.94, 2.54 | 0.080 | 1.36 | 0.81, 2.26 | 0.2 | 26 (1) | 0.60 | 0.03, 4.97 | 0.7 | 0.61 | 0.03, 5.34 | 0.7 |
| Fully vaccinated | 410 (73) | 2.10 | 1.44, 3.09 | <0.001 | 1.90 | 1.28, 2.85 | 0.002 | 482 (29) | 0.96 | 0.32, 4.12 | >0.9 | 0.74 | 0.24, 3.26 | 0.6 |
| **Admitted to hospital for initial COVID-19 infection$** | | | | | | | | | | | | | | |
| No | 1,039 (132) | - | - | - | - | - | - | | | | | | | |
| Yes | 122 (20) | 1.35 | 0.79, 2.21 | 0.3 | 1.34 | 0.77, 2.23 | 0.3 | | | | | | | |

OR = odds ratio, CI = confidence interval, LC = Long COVID. ^All variables were included in the multivariable (adjusted) model. Admission to hospital was not included in the Omicron model, as there were no participants admitted to hospital for their initial COVID-19 infection who reported long COVID.

* self-reported diabetes, cardiovascular disease, HIV, cancer, or asthma.

### partially vaccinated = received some but not all the primary course of a vaccine at time of specimen collection; fully vaccinated = received the primary course of a vaccine at time of specimen collection

$ infection that led to recruitment in the study

Table S6: Factors associated with Long COVID at 3 months, among those infected with Delta and Omicron, excluding those who reported a SARS-CoV-2 infection during follow-up

| | Delta (N=1,076) | | | | | | | Omicron (N=461) | | | | | | |
|---|---|---|---|---|---|---|---|---|---|---|---|---|---|---|
| | | Unadjusted | | | Adjusted^ | | | | Unadjusted | | | Adjusted^ | | |
| | N ($N_{LC}$) | OR | 95% CI | p-value | OR | 95% CI | p-value | N ($N_{LC}$) | OR | 95% CI | p-value | OR | 95% CI | p-value |
| **Age group (years)** | | | | | | | | | | | | | | |
| 0-39 | 597 (64) | - | - | - | - | - | - | 289 (8) | - | - | - | - | - | - |
| 40-54 | 323 (49) | 1.45 | 0.97, 2.17 | 0.068 | 1.13 | 0.74, 1.72 | 0.6 | 116 (13) | 4.43 | 1.82, 11.5 | 0.001 | 4.95 | 1.96, 13.4 | <0.001 |
| 55+ | 156 (29) | 1.90 | 1.16, 3.05 | 0.009 | 1.16 | 0.65, 2.00 | 0.6 | 56 (3) | 1.99 | 0.43, 7.13 | 0.3 | 2.14 | 0.41, 8.64 | 0.3 |
| **Gender** | | | | | | | | | | | | | | |
| Male | 546 (57) | - | - | - | - | - | - | 217 (8) | - | - | - | - | - | - |
| Female | 530 (84) | 1.62 | 1.13, 2.32 | 0.009 | 1.65 | 1.15, 2.39 | 0.007 | 244 (16) | 1.83 | 0.79, 4.60 | 0.2 | 2.04 | 0.86, 5.23 | 0.12 |
| **Chronic health condition*** | | | | | | | | | | | | | | |
| No | 934 (108) | - | - | - | - | - | - | 422 (21) | - | - | - | - | - | - |
| Yes | 142 (33) | 2.32 | 1.48, 3.56 | <0.001 | 1.90 | 1.15, 3.11 | 0.011 | 39 (3) | 1.59 | 0.36, 4.90 | 0.5 | 1.23 | 0.26, 4.42 | 0.8 |
| **COVID-19 vaccination status[#]** | | | | | | | | | | | | | | |
| Not vaccinated | 521 (48) | - | - | - | - | - | - | 43 (3) | - | - | - | - | - | - |
| Partially vaccinated | 182 (26) | 1.64 | 0.97, 2.72 | 0.057 | 1.48 | 0.86, 2.49 | 0.15 | 25 (1) | 0.56 | 0.03, 4.62 | 0.6 | 0.48 | 0.02, 4.24 | 0.5 |
| Fully vaccinated | 373 (67) | 2.16 | 1.45, 3.22 | <0.001 | 2.00 | 1.32, 3.04 | 0.001 | 393 (20) | 0.71 | 0.23, 3.13 | 0.6 | 0.49 | 0.15, 2.24 | 0.3 |

| | | | | | | | |
|---|---|---|---|---|---|---|---|
| **Admitted to hospital for initial COVID-19 infection[$]** | | | | | | | |
| No | 966 (122) | - | - | - | - | - | - |
| Yes | 110 (19) | 1.44 | 0.83, 2.40 | 0.2 | 1.47 | 0.83, 2.49 | 0.2 |

OR = odds ratio, CI = confidence interval, LC = Long COVID. ^All variables were included in the multivariable (adjusted) model. Admission to hospital was not included in the Omicron model, as there were no participants admitted to hospital for their initial COVID-19 infection who reported long COVID.

* self-reported diabetes, cardiovascular disease, HIV, cancer, or asthma.

### partially vaccinated = received some but not all the primary course of a vaccine at time of specimen collection; fully vaccinated = received the primary course of a vaccine at time of specimen collection

$ infection that led to recruitment in the study

Supplementary File 1: List of GISAID IDs used in the phylodynamic analysis

https://unimelbcloud-my.sharepoint.com/:t:/r/personal/wytamma_wirth_unimelb_edu_au/Documents/MDU/CPG/Nepal-COVID-19-2024/GISAID.txt?csf=1&web=1&e=VjV4Nv
